# A multicentre point prevalence survey of patterns and quality of antibiotic prescribing in Indonesian hospitals

**DOI:** 10.1101/2021.03.01.21252506

**Authors:** Ralalicia Limato, Erni J. Nelwan, Manzilina Mudia, Justin de Brabander, Helio Guterres, Enty Enty, Ifael Y. Mauleti, Maria Mayasari, Iman Firmansyah, May Hizrani, Raph L. Hamers

## Abstract

**Synopsis:** *Background:* Antibiotic misuse and overuse are a major driver of antimicrobial resistance, but systematic data in Indonesia are scarce.

*Objectives:* To evaluate patterns and quality indicators of antibiotic prescribing in six acute-care hospitals in Jakarta, Indonesia.

*Methods:* We conducted a hospital-wide point prevalence survey (PPS) between March and August 2019, using Global-PPS and WHO-PPS protocols. The analysis focused on antibacterials for systemic use (antibiotics).

*Results:* Of 1602 inpatients, 993 (62.0%) received ≥1 antimicrobials. Of 1666 antimicrobial prescriptions, 1273 (76.4%) were antibiotics. Most common reasons for prescribing were pneumonia (27.7%), skin and soft tissue infections (8.3%), gastrointestinal prophylaxis (7.9%), and gastrointestinal infections (5.4%). The most common indication was community-acquired infection (42.6%), followed by surgical prophylaxis (22.6%), hospital-acquired infection (18.5%), medical prophylaxis (9.6%), unknown (4.6%) and other indications (2.1%). The most prescribed antibiotic classes were third-generation cephalosporins (44.3%), fluoroquinolones (13.5%), carbapenems (7.4%), penicillins with B-lactamase inhibitor (6.8%) and aminoglycosides (6.0%). According to the WHO AWaRe classification, Watch antibiotics accounted for 67.4%, followed by 28.0% Access, 2.4% Reserve, and 2.2% Unclassified. Reason for prescribing, stop/review date and planned duration were poorly documented. Hospital antibiotic guidelines were not available for 28.1% of prescriptions, and guideline compliance was 52.2% (478/915). Parenteral administration was high (85.1%). Targeted (non-empirical) prescriptions comprised 8.1% (44/542) for community-acquired infections and 26.8% (63/235) for hospital-acquired infections.

*Conclusions:* Our data indicate a high rate of parenteral, empiric use of broad-spectrum antibiotics in Indonesian hospitals, coupled with poor documentation and guideline adherence. The findings suggest important areas for antimicrobial stewardship interventions.

## Introduction

Drug-resistant infections have been estimated to account for 700,000 deaths per year globally, cumulating to 10 million by 2050, higher than cancer (8.2 million) or diabetes (1.5 million) combined.^1^ The overuse and misuse of antimicrobial agents has been well-recognised as one of the key drivers of emerging antimicrobial resistance (AMR),^2,3^ with antimicrobial consumption projected to rise further globally.^4^ In response to the emerging public health crisis of AMR, the World Health Organisation (WHO) has launched a global action plan, including strategies for surveillance and mitigation of antimicrobial overuse.^5^

Indonesia, a populous (271 million) and diverse middle-income country, is potentially an AMR hotspot, due to high infectious disease burdens, liberal antibiotic practices, and fragile health systems.^6,7^ The Indonesian government is increasingly supporting antimicrobial stewardship (AMS), through the national action plan on AMR launched in 2014^8,9^, and as part of hospital accreditation.^10^ However, inappropriate or unnecessary antibiotic prescribing is believed to be widespread, although systematic data are lacking to adequately inform AMS policies.

In global datasets reporting point prevalence surveys (PPS) of antibiotic use in hospitals^11–12^, low and middle-income countries (LMIC) remain underrepresented to date.^13,14^ The recently introduced WHO AWaRe (Access, Watch, Reserve) antibiotic classification framework, based on accessibility versus AMR potential, is a useful metric to provide an indication of the appropriateness of antibiotic consumption.^15,16^

We here report the results of a hospital-wide PPS that evaluated patterns and quality indicators of antibiotic use in six acute-care, general hospitals in Jakarta, the capital city of Indonesia, for community and hospital acquired infections and medical and surgical prophylaxis, by hospital, ward type, indication and diagnosis.

## Patients and Methods

### Study design and population

We conducted a hospital-wide PPS of antimicrobial use in a purposive sample of six hospitals across Jakarta, between March and August 2019. We followed Global PPS (2018)^17^ and WHO (2019)^11^ protocols. Briefly, PPS is a “snap-shot” survey to collate medical record data on antimicrobial prescriptions in hospitalized patients. Eligible patients were all hospitalised patients who received ≥1 active antimicrobial by 8 a.m. on the survey day or surgical prophylaxis ≤24 hours prior to the survey. We excluded emergency and day-care wards, outpatient clinics, inpatients who were discharged before or admitted after 8 a.m.

The study was approved by the Research Ethics Committee of the Faculty of Medicine of the University of Indonesia (1364/UN2.F1/ETIK/2018) and the Oxford Tropical Research Ethics Committee (559-18). The requirement for individual patient consent was waived. Permission was obtained from the hospital management or research/medical committee in each participating hospital.

### Data collection

Data collection was conducted by 1 or 2 medical doctors from the study team, joined by 1 to 4 hospital staff, who received a one-day training and a data collection form (DCF) completion guideline. Each ward was completely surveyed within one day (to minimise the effect of patient movements) and all wards of a single hospital were surveyed within 4 weeks. If crucial patient data was missing, the responsible nurse or clinician was asked for clarifications. A senior team member (RL, RLH) was available to discuss any data queries and take a final decision. We developed a paper DCF comprising ward, patient and treatment sections, modified from Global-PPS^17^ and WHO-PPS.^11^ De-identified data were checked and entered into a study database. We included systemic antimicrobials coded on the basis of the WHO Anatomical Therapeutic Chemical (ATC) classification system as follows: antibacterials (J01), antimycotics (J02), antifungals (D01BA), antimycobacterials (J04), antivirals (J05), nitroimidazole derivatives (P01AB), intestinal antiinfectives (A07A), and antimalarials (P01B). We recorded the diagnosis/reason for the prescribed antimicrobial (what the clinician aimed at treating or preventing), according to a diagnostic code list^17^. Antimicrobial indications were classified as: 1) community-acquired infection (CAI) if symptoms were present on admission or started <48 hours after admission; 2) hospital-acquired infection (HAI) if symptoms started ≥48 hours after admission; 3) medical prophylaxis; 4) surgical prophylaxis, categorised as single-dose, one-day, or longer than one day; 5) other; 6) unknown. We recorded the following five quality indicators of prescribing: 1) documentation of diagnosis/reason for antimicrobial use, stop/review date, and treatment duration; 2) hospital antibiotic guideline availability and compliance with regards to drug choice (if not available, this item was recorded as “not assessable”); 3) parenteral administration; 4) culture sample taken in therapeutic use; 5) targeted (antibiotic prescribed in response to microbiology results) or empirical treatment.

### Statistical analysis

We used descriptive statistics to summarise the data, expressed as counts or percentages, by hospital, ward type, indication and diagnosis. The analysis focused on antibacterials for systemic use (ATC code J01) (aka “antibiotics”). Antibiotics were reported by drug names, chemical class (according to the 4th level WHO ATC classification) and AWaRe groups. We used RStudio Version 1.3.1093 for all analyses.

## Results

### Hospital characteristics

Hospitals were tertiary (2) or secondary-level (4), government or private (3 each), including one specializing in infectious diseases (**Table 1**). All hospitals had an antibiotic stewardship team, five hospitals had local antibiotic guidelines, and four hospitals were included in the national health insurance scheme. All 238 inpatient wards across the six hospitals were surveyed, including 87 medical, 31 surgical, 95 mixed medical-surgical wards and 25 intensive care units, of which 123 adult, 51 paediatric-neonatal and 64 mixed adult-paediatric-neonatal. On the survey day, a total of 2358 active hospital beds (median 230, range 134-853 per hospital) accounted for 1602 (67.9%) admitted patients (median 149, range 51-625 per hospital), of whom 993 (62.0%) received ≥1 antimicrobials (median 91, range 33-368 per hospital). The proportion of patients receiving one or more antimicrobials ranged from 53.5% to 78.8% across hospitals (**Table 1 and S1**).

**Table 1.** Hospital characteristics.

|  | Total | Hospital 1 | Hospital 2 | Hospital 3 | Hospital 4 | Hospital 5 | Hospital 6 |
| --- | --- | --- | --- | --- | --- | --- | --- |
| Level of health service | - | Secondary | Tertiary | Secondary | Secondary | Tertiary | Secondary |
| Sector | - | Private | Public | Private | Public | Public | Private |
| Teaching hospital | - | Yes | Yes | Yes | No | Yes | No |
| National health insurance scheme<sup>a</sup> | - | No | Yes | Yes | Yes | Yes | No |
| Hospital antibiotic guidelines | - | No | Yes | Yes | Yes | Yes | Yes |
| Inpatient wards<sup>b</sup> | 238 | 19 | 74 | 30 | 14 | 79 | 22 |
| Medical wards | 87 | 4 | 27 | 15 | 6 | 30 | 5 |
| Surgical wards | 31 | 0 | 17 | 2 | 0 | 11 | 1 |
| Mixed medical-surgical wards<sup>c</sup> | 95 | 12 | 19 | 10 | 7 | 32 | 15 |
| Intensive care units | 25 | 3 | 11 | 3 | 1 | 6 | 1 |
| Adult | 123 | 8 | 55 | 13 | 2 | 39 | 6 |
| Paediatric and/or neonatal | 51 | 5 | 12 | 6 | 3 | 21 | 4 |
| Mixed adult-neonatal-paediatric<sup>d</sup> | 64 | 6 | 7 | 11 | 9 | 19 | 12 |
| Inpatient beds | 2358 | 159 | 767 | 300 | 145 | 853 | 134 |
| Admitted patients | 1602 (67.9) | 100 (62.9) | 562 (73.3) | 198 (66.0) | 66 (45.5) | 625 (73.3) | 51 (38.1) |
| Admitted patients on ≥1 antimicrobials | 993 (62.0) | 75 (75.0) | 368 (65.5) | 106 (53.5) | 52 (78.8) | 359 (57.4) | 33 (64.7) |
| Medical ward | 310 (31.2) | 47 (62.7) | 102 (27.7) | 45 (42.5) | 14 (26.9) | 95 (26.5) | 7 (21.2) |
| Surgical ward | 184 (18.5) | 0 (0.0) | 87 (23.6) | 24 (22.6) | 0 (0.0) | 73 (20.3) | 0 (0.0) |
| Mixed medical-surgical ward<sup>c</sup> | 401 (40.4) | 23 (30.7) | 135 (36.7) | 37 (34.9) | 35 (67.3) | 147 (40.9) | 24 (72.7) |
| Intensive care unit | 98 (9.9) | 5 (6.7) | 44 (12.0) | 0 (0.0) | 3 (5.8) | 44 (12.3) | 2 (6.1) |
| Adult ward | 727 (73.2) | 31 (41.3) | 298 (81.0) | 92 (86.8) | 43 (82.7) | 230 (64.0) | 33 (100) |
| Paediatric ward | 163 (16.4) | 13 (17.4) | 58 (15.7) | 8 (7.5) | 3 (5.7) | 81 (22.6) | 0 (0.0) |
| Mixed adult-neonatal-paediatric ward<sup>d</sup> | 103 (10.4) | 31 (41.3) | 12 (3.3) | 6 (5.7) | 6 (11.6) | 48 (13.4) | 0 (0.0) |

Data shown reflect the hospital situation on the survey day, and are expressed as number (percentage), unless otherwise specified.
<sup>a</sup> National health insurance scheme, *Jaminan Kesehatan Nasional (JKN)*.
<sup>b</sup> Includes all inpatients wards in the hospital. Some wards have been further subdivided for the purpose of this survey.
<sup>c</sup> Wards that can admit both medical and surgical patients.
<sup>d</sup> Wards that can admit both adult, paediatric and neonatal patients.

### Characteristics of patients receiving one or more antimicrobials

**Table 2** summarizes patient characteristics. Of 993 patients, 497 (50.1%) were women and 782 (78.8%) were adults, and the median age was 43 years (IQR 22-58.5; range 1 day to 99 years). One or more comorbidities were documented in 48.9% (486) of patients. 368 (37.1%) had undergone surgery and 299 (30.1%) had been hospitalised in the last 90 days, and 145 (14.6%) had been transferred from another hospital.

**Table 2.** Characteristics of patients receiving ≥1 antimicrobials.

|  | **Total**<br>**(n=993)** | **Hospital 1**<br>**(n=75)** | **Hospital 2**<br>**(n=368)** | **Hospital 3**<br>**(n=106)** | **Hospital 4**<br>**(n=52)** | **Hospital 5**<br>**(n=359)** | **Hospital 6**<br>**(n=33)** |
| --- | --- | --- | --- | --- | --- | --- | --- |
| Female | 497 (50.1) | 41 (54.7) | 186 (50.5) | 61 (57.5) | 22 (42.3) | 175 (48.7) | 12 (36.4) |
| Age (median, IQR)<sup>a</sup> | 43 (22-58.5) | 29 (20-55) | 47 (25-60) | 52 (33-66) | 39.5 (28-59) | 37 (8-53) | 51 (28.5-65) |
| <1 month (neonates) | 45 (4.5) | 2 (2.7) | 17 (4.6) | 2 (1.9) | 1 (1.9) | 23 (6.4) | 0 (0.0) |
| 1-23 months | 63 (6.3) | 3 (4.0) | 20 (5.4) | 2 (1.9) | 1 (1.9) | 37 (10.3) | 0 (0.0) |
| 2-17 years | 103 (10.4) | 12 (16.0) | 24 (6.5) | 6 (5.7) | 4 (7.7) | 57 (15.9) | 0 (0.0) |
| 18-29 years | 131 (13.2) | 21 (28.0) | 47 (12.8) | 7 (6.6) | 8 (15.4) | 39 (10.9) | 9 (27.3) |
| 30-39 years | 112 (11.3) | 9 (12.0) | 35 (9.5) | 24 (22.6) | 12 (23.1) | 30 (8.4) | 2 (6.1) |
| 40-49 years | 145 (14.6) | 7 (9.3) | 55 (14.9) | 7 (6.6) | 8 (15.4) | 63 (17.5) | 5 (15.2) |
| ≥50 years | 394 (39.7) | 21 (28.0) | 170 (46.2) | 58 (54.7) | 18 (34.6) | 110 (30.6) | 17 (51.5) |
| National health insurance holder<sup>b</sup> | 743 (74.8) | 0 (0.0) | 329 (89.4) | 24 (22.6) | 50 (96.2) | 340 (94.7) | 0 (0.0) |
| Transfer from other hospital | 145 (14.6) | 2 (2.7) | 70 (19.0) | 2 (1.9) | 10 (19.2) | 57 (15.9) | 4 (12.1) |
| Hospitalisation within 90 days<sup>c</sup> | 299 (30.1) | 12 (16.0) | 86 (23.4) | 20 (18.9) | 20 (38.5) | 151 (42.1) | 10 (30.3) |
| Surgery in the past 90 days<sup>d</sup> | 368 (37.1) | 3 (4.0) | 162 (44.0) | 41 (38.7) | 8 (15.4) | 142 (39.6) | 12 (36.4) |
| Catheter use |  |  |  |  |  |  |  |
| Central vascular | 132 (13.3) | 4 (5.3) | 35 (9.5) | 12 (11.3) | 1 (1.9) | 73 (20.3) | 7 (21.2) |
| Peripheral vascular | 941 (94.8) | 69 (92.0) | 357 (97.0) | 85 (80.2) | 51 (98.1) | 347 (96.7) | 32 (97.0) |
| Urinary | 363 (36.6) | 9 (12.0) | 183 (49.7) | 35 (33.0) | 9 (17.3) | 119 (33.1) | 8 (24.2) |
| Intubation | 65 (6.5) | 3 (4.0) | 21 (5.7) | 7 (6.6) | 1 (1.9) | 30 (8.4) | 3 (9.1) |
| Documented comorbidity | 486 (48.9) | 11 (14.6) | 209 (56.8) | 40 (37.7) | 41 (78.8) | 174 (48.5) | 14 (42.4) |
| Malnutrition | 335 (33.7) | 0 (0.0) | 165 (44.8) | 17 (16.0) | 28 (53.8) | 121 (33.7) | 4 (12.1) |
| Diabetes mellitus | 161 (16.2) | 8 (10.7) | 68 (18.5) | 24 (22.6) | 14 (26.9) | 39 (10.9) | 8 (24.2) |
| Tuberculosis | 120 (12.1) | 3 (4.0) | 37 (10.1) | 7 (6.6) | 24 (46.2) | 44 (12.3) | 5 (15.2) |
| HIV | 44 (4.4) | 3 (4.0) | 10 (2.7) | 2 (1.9) | 13 (25.0) | 16 (4.5) | 0 (0.0) |
| HIV on antiretroviral treatment | 27 (2.7) | 3 (4.0) | 4 (1.1) | 2 (1.9) | 8 (15.4) | 10 (2.8) | 0 (0.0) |
| Chronic obstructive pulmonary disease | 11 (1.1) | 0 (0.0) | 7 (1.9) | 0 (0.0) | 3 (5.8) | 1 (0.3) | 0 (0.0) |
| McCabe score<sup>e</sup> |  |  |  |  |  |  |  |
| Rapidly fatal | 29 (2.9) | 3 (4.0) | 8 (2.2) | 2 (1.9) | 0 (0.0) | 15 (4.2) | 1 (3.0) |
| Ultimately fatal | 217 (21.9) | 3 (4.0) | 89 (24.2) | 32 (30.2) | 2 (3.8) | 84 (23.4) | 7 (21.2) |
| Non-fatal | 746 (75.1) | 69 (92.0) | 270 (73.4) | 72 (67.9) | 50 (96.2) | 260 (72.4) | 25 (75.8) |
| Unknown | 1 (0.1) | 0 (0.0) | 1 (0.1) | 0 (0.0) | 0 (0.0) | 0 (0.0) | 0 (0.0) |
| Prescribed antimicrobial drugs | 1,666 | 114 | 630 | 158 | 98 | 622 | 44 |
| Median (range) per patient | 1 (1-12) | 1 (1-12) | 1 (1-9) | 1 (1-7) | 1 (1-6) | 1 (1-10) | 1 (1-4) |
| 1 | 602 (60.6) | 58 (77.3) | 204 (55.4) | 74 (69.8) | 32 (61.5) | 209 (58.2) | 25 (75.8) |
| 2 | 254 (25.6) | 11 (14.7) | 108 (29.3) | 22 (20.8) | 8 (15.4) | 99 (27.6) | 6 (18.2) |
| ≥3 | 137 (13.8) | 6 (8.0) | 56 (15.2) | 10 (9.4) | 12 (23.1) | 51 (14.2) | 2 (6.1) |

Data shown reflect the hospital situation on the survey day, and are expressed as number (percentage), unless otherwise specified
<sup>a</sup> Median (IQR) age was 47 (28-60) years for adults, 7 (2-11) months for children <2 years, and 8 (3-14) days for neonates
<sup>b</sup> Jaminan Kesehatan Nasional (JKN); Unknown for 4 (0.4%) participants.
<sup>c</sup> Before the current admission, the patient has been hospitalized within 90 days before the survey date.
<sup>d</sup> The patient underwent surgery in the past 90 days before the survey date, including surgery prior to and during the current admission
<sup>e</sup> McCabe score is a simple subjective method to assess underlying illness severity and classify patients according to a prognosis of rapidly fatal (<1 year), ultimately fatal (1–4 years) and non-fatal (>5 years) (Reilly et al. J Infect Prev 2016;17(3):127–129).

The 993 patients receiving one or more antimicrobials accounted for a total of 1666 active antimicrobial prescriptions (median 1 per patient, range 1-12), with 60.6% (602) receiving one antimicrobial agent, 25.6% (254) two, and 13.8% (137) three or more. Antimicrobial use was highest in ICU (86.8%, 132/152), followed by surgical wards (66.0%, 184/293), mixed medical-surgical wards (65.0%, 401/622) and medical wards (51.4%, 310/569) (**Table S1**). Concomitant use of ≥2 antimicrobials was more frequent in ICU (59.1%, 58/98) than non-ICU (37.2%, 333/895) (**Table 2**).

### Antimicrobial agents prescribed

Of all 1666 antimicrobial prescriptions, 76.4% (1273) were antibacterials, followed by 11.4% (197) antimycobacterials, 4.3% (72) antivirals, 3.7% (62) antimycotics, 2.6% (43) intestinal antiinfectives, 0.8% (13) antimalarials, and 0.4% (6) nitroimidazole derivatives. The 1273 prescribed antibiotics (J01) included 46 different agents. The five most prescribed antibiotics were ceftriaxone (26.8%, 341), levofloxacin (10.7%, 137), metronidazole (7.1%, 91), meropenem (6.4%, 82) and cefotaxime (5.6%, 71) –accounting for 56.5% (720) of prescriptions (**Table S2**). The five most prescribed antibiotic classes were third-generation cephalosporins (44.3%, 565), fluoroquinolones (13.5%, 172), carbapenems (7.4%, 94), penicillins with B-lactamase inhibitor (6.8%, 86) and aminoglycosides (6.0%, 76) (**Figure 1**) –accounting for 78.0% (993) of prescriptions.

**Figure 1.**
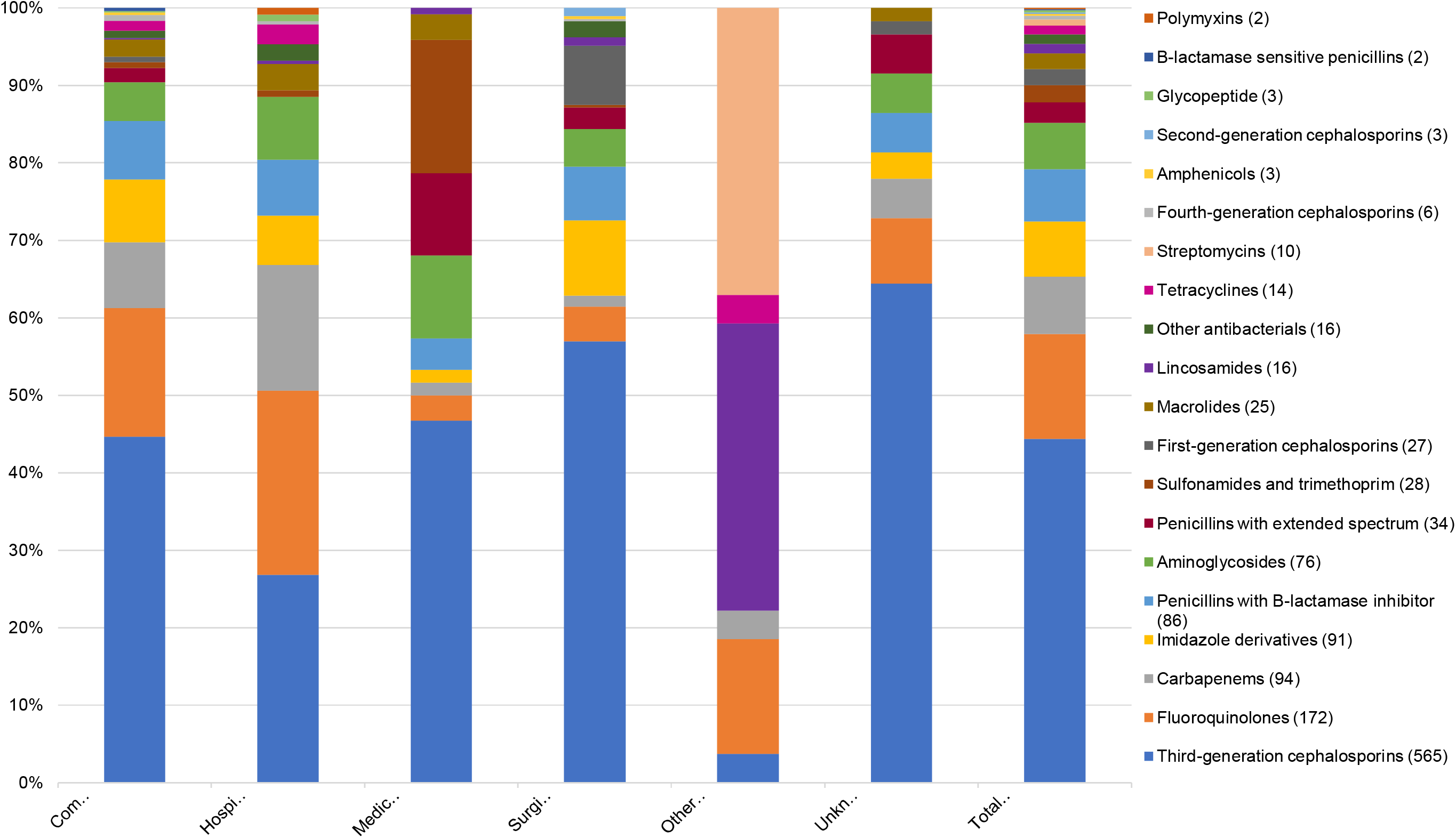
Systemic antibiotic use by antibiotic class, by indication.

### Reasons for antibiotic prescribing

The full list of diagnosis/reasons for all 1666 antimicrobial prescriptions is included in the data supplement (**Table S3)**. Among all 1273 antibiotic prescriptions (J01), the most common diagnosis/reasons were pneumonia (27.7%, 353), skin and soft tissue infections (8.3%, 106), gastrointestinal prophylaxis (7.9%, 101), and gastrointestinal infections (5.4%, 69) –accounting for 49.4% (629) of all prescriptions (**Table 3** and **S4**). Ceftriaxone and levofloxacin were mainly used to treat pneumonia and gastrointestinal infections. Metronidazole was mainly used for skin and soft tissue infections and intra-abdominal infections, and for gastrointestinal prophylaxis. Meropenem was mainly used for pneumonia and sepsis. Cefotaxime was mainly used for pneumonia and for gastrointestinal prophylaxis.

**Table 3.** Most common diagnosis for systemic antibiotic use.

| Diagnosis | Total (n=1,273) |
| --- | --- |
| Pneumonia or lower respiratory tract infection | 353 (27.7) |
| Skin and soft tissue infection <sup>a</sup> | 106 (8.3) |
| Prophylaxis for gastrointestinal infections <sup>b</sup> | 101 (7.9) |
| Gastrointestinal infection | 69 (5.4) |
| Prophylaxis for bone and joint infection <sup>c</sup> | 58 (4.6) |
| Prophylaxis for obstetrics or gynaecological infection | 58 (4.6) |
| Intra-abdominal infection <sup>d</sup> | 57 (4.5) |
| Unknown reason | 47 (3.7) |
| Sepsis | 46 (3.6) |
| Prophylaxis for urinary tract infection (surgery or recurrent infection) | 42 (3.3) |
| Other diagnosis <sup>e</sup> | 35 (2.7) |
| Ear, nose, throat infection <sup>f</sup> | 30 (2.4) |
| Upper urinary tract infection <sup>g</sup> | 29 (2.3) |
| Central nervous system infection | 26 (2.0) |
| Medical prophylaxis for new-born risk factors | 25 (2.0) |

<sup>a</sup> Including cellulitis, wound including surgical site infections, deep soft tissue not involving bone (e.g. infected pressure or diabetic ulcers, abscess).
<sup>b</sup> Including prophylaxis for surgery of the gastrointestinal tract, liver, or biliary tree, and prophylaxis in patients with neutropenia or hepatic failure.
<sup>c</sup> Including prophylaxis for surgical site infections, for plastic or orthopaedic surgery (bone or joint).
<sup>d</sup> Including hepatobiliary, intra-abdominal abscess, etc.
<sup>e</sup> Antibiotic prescribed with documentation for which there is no above diagnosis group.
<sup>f</sup> Including mouth, sinuses, larynx.
<sup>g</sup> Including catheter related urinary tract infection, pyelonephritis.

### Indications for antibiotic prescribing

Among all 1273 antibiotic prescriptions (J01), the most common indication was CAI (42.6%, 542), followed by surgical prophylaxis (22.6%, 288), HAI (18.5%, 235), medical prophylaxis (9.6%, 122), unknown (4.6%, 59), and other (2.1%, 27) (**Table S5**)

Among CAI (542), the top-5 diagnosis/reasons were pneumonia (42.6%, 231), skin and soft tissue infection (14.2%, 77), gastrointestinal infection (12.2%, 66), sepsis (5.5%, 30), and intra-abdominal infection (5.4%, 29) (**Table S4**). The five most common antibiotics for CAI were ceftriaxone (32.8%, 178), levofloxacin (13.5%, 73), metronidazole (8.1%, 44), meropenem (7.7%, 42), and ampicillin sulbactam (5.4%, 29) –accounting for 67.5% (366) of prescriptions (**Table S6**).

The most common HAI was hospital-acquired pneumonia (including other HAI) (70.2%, 165), followed by intervention-related infections (including catheter-related blood stream infection, ventilator-associated pneumonia, catheter-related urinary tract infection) (19%, 35), post-operative surgical site infection (13.6%, 32), infection present on admission from another hospital (0.85%, 2) or long-term care facility (0.4%, 1); no *C. difficile*-associated diarrhoea (HAI3) was documented (**Table S7**). The six most common antibiotics for HAI were levofloxacin (18.7%, 44), meropenem (13.6%, 32), ceftriaxone (9.8%, 23), amikacin (6.4%, 15), metronidazole and ceftazidime (6%, 14 each) –accounting for 60.4% (142) of prescriptions (**Table S6**).

Among medical prophylaxis (122), the top-5 diagnosis/reasons were neonatal (20.5%, 25), general (19.7%, 24), gastrointestinal (17.2%, 21), respiratory (15.6%, 19), and unknown (8.2%, 10) (**Table S4**). The five most common antibiotics for medical prophylaxis were ceftriaxone (28.7%, 35), cotrimoxazole (17.2%, 21), gentamicin (10.7%, 13), cefotaxime (8.2%, 10), ampicillin (7.4%, 9) – accounting for 72.1% (88) of prescriptions (**Table S6**).

Among surgical prophylaxis (288), the top-5 diagnosis/reasons were gastrointestinal (27.8%, 80), obstetrics/gynaecology (20.1%, 58), bone and joint (17%, 49), urinary tract (12.8%, 37), central nervous system and ear-nose-throat (7.3%, 21 each). The most common antibiotics for surgical prophylaxis were ceftriaxone (26.4%, 76), cefixime (11.5%, 33), cefoperazone (11.1%, 32), metronidazole (9.7%, 28), cefazolin (6.2%, 18) –accounting for 64.9% (187) of prescriptions. Notably, duration of surgical prophylaxis was longer than one day for 76% (219) of prescriptions, whereas 15.0% (43) was single-dose and 9.0% (26) was for one day.

### Antibiotic use based on AWaRe groups

**Figure 2** summarizes AWaRe groups. Of all 1273 antibiotic prescriptions (J01), 67.4% (858) were Watch antibiotics, followed by 28.0% (356) Access, 2.4% (31) Reserve, and 2.2% (28) Unclassified. This pattern was similar across indications and ward types. Of note, Watch antibiotics were commonly prescribed for the most frequent diagnosis, i.e. pneumonia (34.4%, 295), gastrointestinal infection (6.6%, 57), and skin and soft tissue infection (5.8%, 50). Watch antibiotics (858) comprised 45.1% (387) of CAI, 20.9% (179) of surgical prophylaxis, 19.5% (167) of HAI, and 7.2% (62) of medical prophylaxis. The five most used Watch antibiotics were ceftriaxone (39.7%, 341), levofloxacin (15.8%, 136), meropenem (9.6%, 82), cefotaxime (8.3%, 71), and cefoperazone (6%, 52) (**Table S7**).

**Figure 2.**
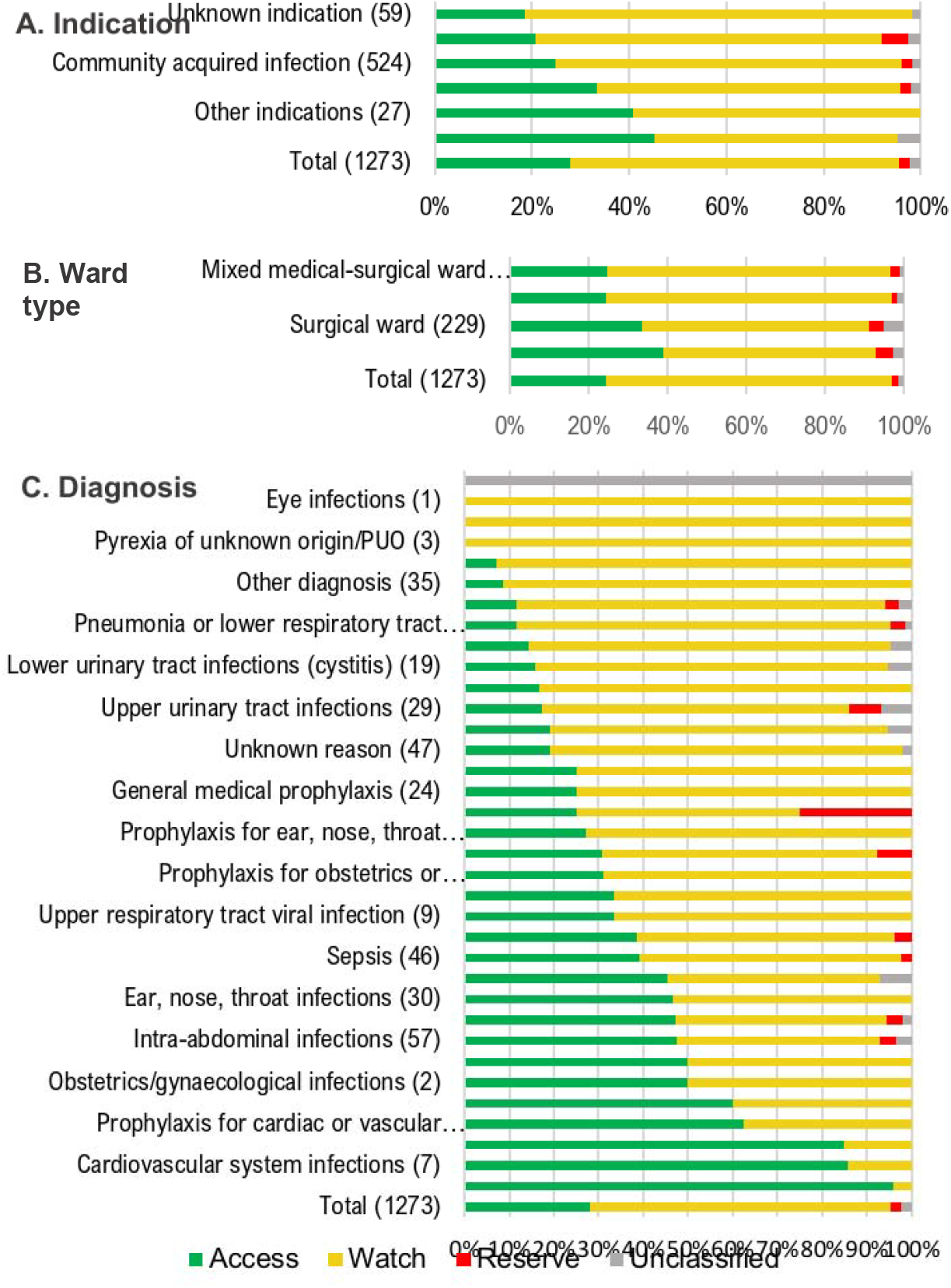
Systemic antibiotic use by AWaRe classification.

Access antibiotics (356) comprised 37.6% (134) of CAI, 27% (96) of surgical prophylaxis, 15.4% (55) of medical prophylaxis, and 13.8% (49) of HAI. The five most used Access antibiotics were metronidazole (25.3%, 90), ampicillin-sulbactam (12.4%, 44), gentamicin (11.8%, 42), amikacin (9%, 32) and amoxicillin-clavulanic acid (8.7%, 31) (**Table S7, Figure 3**).

**Figure 3.**
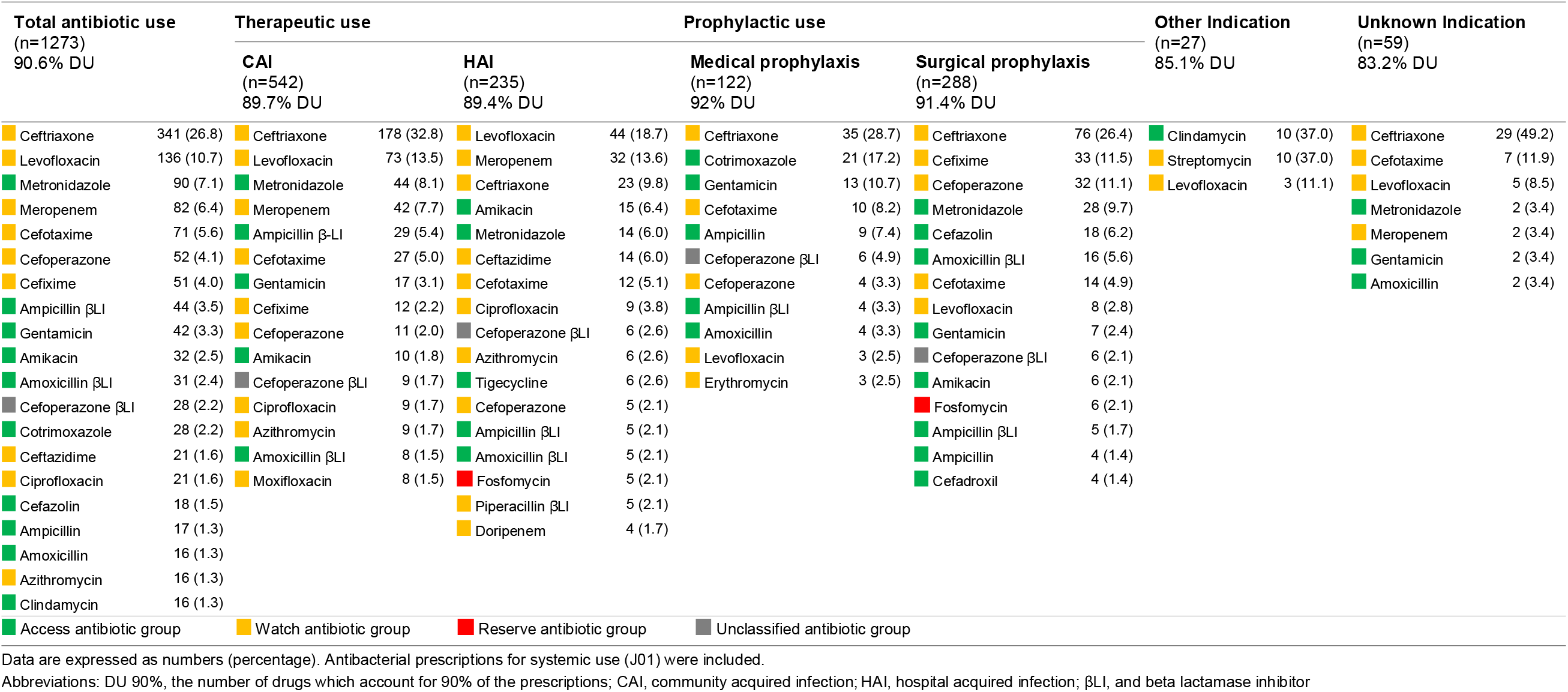
Systemic antibiotic use by indication based on AWaRe classification.

Reserve antibiotics were uncommon (31), and included fosfomycin (15, 48.4%), tigecycline (13, 41.9%), colistin (1, 3.2%), and linezolid (1, 3.2%).

### Quality indicators of antibiotic use (J01)

#### Documentation of antibiotic plan

Reason for prescribing was documented for 63.5% (808/1273) of all prescriptions, with substantial variation between hospitals, indications and ward types (**Table 4 and S9**). Documentation of diagnosis/reason was better for therapeutic use (546/777, 70.3%) than prophylactic use (230/410, 56.0%), and in ICU (75.6%, 118/156) than non-ICU (61.8%, 690/1117). Stop/review date (15.2%, 194/1273) and planned treatment duration (9.8%, 125/1273) was poorly documented overall, across indications and ward types (**Table 4 and S9**).

**Table 4.** Quality indicators for antibiotic prescribing, by indication.

| Quality indicators | Total<br>(n=1,273) | Therapeutic use<br>(n=777) |  |  |  | Prophylactic use<br>(n=410) |  | Other<br>indication<sup>a</sup><br>(n=27) | Unknown<br>indication<br>(n=59) |
| --- | --- | --- | --- | --- | --- | --- | --- | --- | --- |
|  |  | CAI empirical<br>(n=498) | CAI targeted<br>(n=44) | HAI empirical<br>(n=172) | HAI targeted<br>(n=63) | Medical<br>prophylaxis<br>(n=122) | Surgical<br>prophylaxis<br>(n=288) |  |  |
| Reason documented | 808 (63.5) | 321 (64.5) | 35 (79.5) | 145 (84.3) | 45 (71.4) | 58 (47.5) | 172 (59.7) | 21 (77.8) | 11 (18.6) |
| Stop/review date documented | 194 (15.2) | 69 (13.9) | 7 (15.9) | 43 (25.0) | 18 (28.6) | 15 (12.3) | 38 (13.2) | 2 (7.4) | 2 (3.4) |
| Treatment duration documented | 125 (9.8) | 35 (7.0) | 5 (11.4) | 31 (18.0) | 12 (19.0) | 3 (2.5) | 35 (12.2) | 0 (0.0) | 4 (6.8) |
| Guideline compliance |  |  |  |  |  |  |  |  |  |
| Yes | 478 (37.5) | 223 (44.8) | 17 (38.6) | 79 (45.9) | 33 (52.4) | 23 (18.9) | 82 (28.5) | 21 (77.8) | 0 (0.0) |
| No | 378 (29.7) | 136 (27.3) | 16 (36.4) | 70 (40.7) | 20 (31.7) | 6 (4.9) | 124 (43.1) | 6 (22.2) | 0 (0.0) |
| Not assessable<sup>b</sup> | 358 (28.1) | 139 (27.9) | 11 (25.0) | 23 (13.4) | 10 (15.9) | 93 (76.2) | 82 (28.5) | 0 (0.0) | 0 (0.0) |
| Indication unknown | 59 (4.6) | 0 (0.0) | 0 (0.0) | 0 (0.0) | 0 (0.0) | 0 (0.0) | 0 (0.0) | 0 (0.0) | 59 (100.0) |
| Route of administration |  |  |  |  |  |  |  |  |  |
| Parenteral (IV) | 1,084 (85.2) | 444 (89.2) | 42 (95.5) | 150 (87.2) | 58 (92.1) | 92 (75.4) | 237 (82.3) | 10 (37.0) | 51 (86.4) |
| Oral | 183 (14.38) | 52 (10.44) | 2 (4.55) | 22 (12.79) | 5 (7.94) | 30 (24.59) | 51 (17.71) | 13 (48.15) | 8 (13.56) |
| IV-oral switch | 48 (26.2) | 6 (11.5) | 1 (50.0) | 9 (40.9) | 1 (20.0) | 1 (3.3) | 28 (54.9) | 0 (0.0) | 2 (25.0) |
| Other | 6 (0.47) | 2 (0.40) | 0 (0.00) | 0 (0.00) | 0 (0.00) | 0 (0.00) | 0 (0.00) | 4 (14.81) | 0 (0.00) |
| Culture sample taken<sup>c</sup> | 344 (44.3) | 125 (25.1) | 44 (100) | 112 (65.1) | 63 (100) | - | - | - | - |

Abbreviations: CAI, community acquired infection; HAI, hospital acquired infection; IV, intravenous
<sup>a</sup> Other indication included antibiotics prescribed for neurotoxoplasmosis, pulmonary tuberculosis, and as motility agent.
<sup>b</sup> Hospital antibiotic guidelines were not available to assess compliance.
<sup>c</sup> Only applicable to therapeutic use.

#### Hospital guideline availability and compliance

Local antibiotic guidelines were not available for 28.1% (358/1273) of antibiotic prescriptions; and, of note, for 76.2% (93/122) of prescriptions for medical prophylaxis. Guideline compliance was 52.2% (478/915) overall, 44.8% (223/498) for empirical CAI treatment, 45.9% (79/172) for empirical HAI treatment, 28.5% (82/288) for surgical prophylaxis and 18.9% (23/122) for medical prophylaxis (**Table 4**). Guideline compliance was similar across ward types (**Table S9**).

#### Parenteral use

85.1% (1084/1273) of prescriptions were parenterally administered, including 88.5% (208/235) for HAI, 89.5%% (486/542) for CAI, 75.4% (92/122) for medical prophylaxis and 82.3% (237/288) for surgical prophylaxis (Table 4). Parenteral use was higher in ICU (97.4%, 152/156) than non-ICU (83.4%, 932/1117) (**Table S9**).

#### Culture samples taken

Among 619 patients with ≥1 antibiotic for therapeutic use, 48.8% (302) had one or more samples taken for bacterial culture (total 831 samples, median 2, range 1-25 per patient). Among the top-5 diagnosis/reasons for prescribing, one or more culture sample were taken in 44.8% (154/353) of pneumonias, 51.9% (55/106) of skin and soft tissue infections, 66.7% (38/57) of intra-abdominal infections, 52.2% (24/46) of sepsis, and 75.9% (22/29) of upper UTI. Blood cultures were taken in 44.4% (88/353) of pneumonias, 45.6% (26/57) of intra-abdominal infections, 58.6% (17/29) of upper UTI, and 95.8% (23/24) of sepsis. Sputum cultures were taken in 26.9% (95/353) of pneumonias. Urine cultures were taken in 72.4% (21/29) of upper UTI (**Table S10**).

#### Targeted antibiotic treatment

Treatment was targeted in 8.1% (44/542) of CAI and 26.8% (63/235) of HAI (**Table 4**); 13.0% (46/353) of pneumonias, 15.8% (9/57) of intra-abdominal infections, 44.8% (13/29) of upper UTI, and 13% (6/46) of sepsis. Targeted treatment was more common in ICU (19.9%, 31/156) than in non-ICU (6.8%, 76/1117) (**Table S9**).

## Discussion

This was the first contemporary hospital-wide survey in Indonesia that systematically evaluated patterns and quality of antibiotic prescribing, using the recommended PPS methodology.^11,17^ We demonstrated the feasibility of PPS in this low-resource setting, and generated useful data to guide local AMS programmes. Our survey found a high proportion (62%) of antimicrobial use among hospitalized patients, ranging from 53.5% to 78.8% across hospitals in Jakarta, with the highest use in ICU (86.8%) as expected. Antibiotic use was substantially higher than reported in global PPS datasets, which were dominated by data from high-income countries in Europe, North America and Asia^13,14^ and which highlighted that antimicrobial use was significantly higher in non-European hospitals compared with European hospitals.^14^

Consistent with other surveys in Asia^18,19^ and globally^13^, the predominant reason for antibiotic prescription in Jakarta hospitals were lower respiratory tract infections. In our survey, the mostly used antibiotic classes were third-generation cephalosporines (mainly ceftriaxone), fluoroquinolones (mainly levofloxacin), and carbapenems (mainly meropenem), all predominantly used for pneumonia, among several other diagnosis. Ceftriaxone was the most used antibiotic across all major indications (i.e. CAI, HAI, surgical and medical prophylaxis). These findings are consistent with the widespread use of broad-spectrum antibiotics, predominantly third-generation cephalosporins and fluoroquinolones, in Indonesia^20^, other Asian countries^18,19,21–23^ and globally^13,14^, which may suggest that at least a proportion of these prescriptions are unnecessary or inappropriate. Moreover, the empirical use of meropenem for CAI and HAI represented nearly 10% of all antibiotics for therapeutic use. This could partially be explained by high rates of AMR, particularly in common Gram-negative organisms, in Indonesian hospitals.^24^ However, targeted prescribing for CAI (8%) and HAI (27%) was low in comparison to a global study (12-27% and 20-44%, respectively)^13^, suggesting underutilization of microbiological diagnostics as well as overuse of broad-spectrum antibiotics.

Antibiotic prescriptions for HAI (18.5% of total), predominantly for pneumonia but also intervention-related and post-operative surgical site infections, were comparable to recent surveys in India (19%)^19^ and Thailand (34%)^18^, but considerably higher than in reports from high-income settings, e.g. ECDC survey (6%)^25^ and the GLOBAL-PPS survey (8.4%).^13^ These data confirm the significantly higher burden of HAI in LMIC compared to high-income countries.

A high proportion of antibiotic prescriptions were for surgical (23%) and medical prophylaxis (10%), for a range of indications. Prophylactic prescribing was unusually high for gastrointestinal infections. Prolonged (>1 day) surgical prophylaxis was very common (76%) in our survey, as has also been observed in other countries in Asia (Pakistan 97%^21^, India 77%^19^, Thailand 90%^18^) as well as in Europe^13,25^. Prolonged antibiotic prophylaxis for more than 24h for most surgical indications does not prevent development of postoperative infections, compared with <24h, but increases the risk of AMR and side-effects.^26^ Further research is warranted to explain the reasons for these patterns.

We investigated five basic quality indicators, which could be used to set benchmarks for quality improvement of antibiotic use^27^ and AMS programmes.^28^ Documentation of the reason of prescribing (64%) was lower than reported across studies in Europe, Asia, Africa, and America (70-85%).^13,29^ Stop or review date was poorly documented (15%) across indications and ward types. Post-prescription review of a prescribed antimicrobial within 48-72h of the initial order, ensures appropriate choice and route of administration, optimal de-escalation (IV-oral switch) practices, and prevents unnecessarily long antibiotic courses. The high (85%) proportion of parenteral route of administration, coupled with high rates of empirical therapy and suboptimal use of microbiological cultures, suggests lack of de-escalation protocols in the participating hospitals. Pro-active IV to oral switching policies can reduce catheter-related complications, health-care costs, and duration of hospital stays, and is recognized as a key metric for AMS processes ^30^.

Hospital guidelines were not available for 28% of antibiotic prescriptions, including for 76% of prescriptions for medical prophylaxis. A systematic review and meta-analysis showed that guideline-adherent empirical therapy was associated with a relative risk reduction for mortality of 35%^30^. The reason for poor guideline compliance is uncertain and probably multifactorial, including local resistance patterns, ineffective guideline dissemination, and clinical uncertainty with fear of treatment failure. Our findings should trigger further detailed investigations at hospital and country level.

The WHO AWaRe framework offers an attractive metric for LMIC in the absence of validated quality indicators for antibiotic appropriateness^15,31,32^, and includes a >60% national target of total antibiotic consumption in the Access category by 2023^33^. However, a recent assessment of antibiotic consumption data from 76 countries in 2000-2015 found that the global per-capita consumption of Watch antibiotics increased by 90.9%, compared with an increase of 26.2% in Access antibiotics, with disproportionate increases in Watch antibiotic consumption in LMIC (165% compared with 27.9% in high-income countries)^16^. Although Indonesia national-level data have not been included in the AWaRE reports to date^33^, our survey found hospital consumption of Access antibiotics at 28% to be far below the 60% target, mostly driven by ceftriaxone and levofloxacin use for CAI and HAI. Although these findings could partially be explained by the national health insurance scheme which determines available antibiotics based on the national formulary^34^, they also highlight significant challenges for AMS.

The limitations of this study are inherent to the cross-sectional “snap-shot” design. As we used a convenient hospital sample, data are not representative for all hospitals in Jakarta or Indonesia, urging caution in extrapolating the observed patterns. Antibiotic use patterns can be influenced by many factors, e.g. patient case mix, prevalence of different types of infections, AMR patterns, institutional factors, among others.

In conclusion, we observed high levels of parenteral, empiric use of broad-spectrum antibiotics in Indonesian hospitals, and inadequate performance on key quality indicators of prescribing. Despite important progress in AMS, supported by national policies^9,10^ and guidelines^35^, the study findings highlighted the need to strengthen AMS to increase use of narrower-spectrum antibiotics through culture-guided targeted treatment and hospital guideline compliance. Further research is needed to understand the complex drivers of antibiotic prescribing, and to develop context-specific and feasible quality improvement strategies to strengthen existing AMS programmes.

## Data Availability

After publication, the data will be made available to others on reasonable requests to the corresponding author, including a detailed research proposal, study objectives and statistical analysis plan. Deidentified participant data will be provided after written approval from the corresponding author and data contributors.

## Acknowledgements

We thank the management, research/medical committees and clinical staff of the participating hospitals for their support, and Reinout van Crevel and Rogier van Doorn for useful feedback on the manuscript.

## EXPLAIN study group

Ralalicia Limato, Erni J. Nelwan, Manzilina Mudia, Helio Guterres, Enty Enty, Ifael Y. Mauleti, Maria Mayasari, Iman Firmansyah, May Hizrani, Raph L. Hamers, Anis Karuniawati, Prof Taralan Tambunan, Prof Amin Soebandrio, Prof Henri Verbrugh, Decy Subekti, Iqbal Elyazar, Mutia Rahardjani, Fitria Wulandari, Prof Reinout van Crevel, Rogier van Doorn, Vu Thi Lan Huong, Nga Do Ti Thuy, Sonia Lewycka, Prof Alex Broom.

## Funding

This work was funded by the Wellcome Trust, UK (106680/Z/14/Z)

## Transparency declarations

### Conflicts of interest

The authors declare no competing interests.

### Author contributions

EJN and RLH conceived the idea for the study and are the principal investigators. RLH obtained the funding. RL, EJN and RLH designed the study protocol. MM, HG, EE, IYM, MM, IF, and MH collected and verified the data, overseen by RL. RL and MM curated the database, did the analysis and had full access to all study data, supervised by EJN and RLH. RL, MM and RLH drafted the paper. All authors critically revised the manuscript and all authors gave approval for the final version to be published.

## Notes

### Competing Interest Statement

The authors have declared no competing interest.

