## Supplemental File for "A multicentre point prevalence survey of patterns and quality of antibiotic prescribing in Indonesian hospitals"

**Data supplement**

**Table S1. Systemic antimicrobial use, by hospital and by ward type**

| **Wards** | **Total**  **(n=238)** | **Hospital 1**  **(n=19)** | **Hospital 2**  **(n=74)** | **Hospital 3**  **(n=30)** | **Hospital 4**  **(n=14)** | **Hospital 5**  **(n=79)** | **Hospital 6**  **(n=22)** |
| --- | --- | --- | --- | --- | --- | --- | --- |
| Active bed | 2358 | 159 | 767 | 300 | 145 | 853 | 134 |
| Admitted patients | 1602 (67.9) | 100 (62.9) | 562 (73.3) | 198 (66.0) | 66 (45.5) | 625 (73.3) | 51 (38.1) |
| Patients on ≥1 antimicrobial | 993 (62.0) | 75 (75.0) | 368 (65.5) | 106 (53.5) | 52 (78.8) | 359 (57.4) | 33 (64.7) |
| **By ward type** |  |  |  |  |  |  |  |
| Medical ward |  |  |  |  |  |  |  |
| Admitted patients | 569 | 57 | 167 | 106 | 19 | 210 | 10 |
| Patients on ≥1 antimicrobial | 310 (54.5) | 47 (82.5) | 102 (61.1) | 45 (42.5) | 14 (73.7) | 95 (45.2) | 7 (70.0) |
| Surgical ward |  |  |  |  |  |  |  |
| Admitted patients | 293 | 0 | 119 | 32 | 0 | 141 | 1 |
| Patients on ≥1 antimicrobial | 184 (62.8) | 0 (0.0) | 87 (73.1) | 24 (75.0) | 0 (0.0) | 73 (51.8) | 0 (0.0) |
| Mixed medical-surgical ward |  |  |  |  |  |  |  |
| Admitted patients | 622 | 38 | 215 | 60 | 44 | 227 | 38 |
| Patients on ≥1 antimicrobial | 401 (64.5) | 23 (60.5) | 135 (62.8) | 37 (61.7) | 35 (79.5) | 147 (64.8) | 24 (63.2) |
| ICU |  |  |  |  |  |  |  |
| Admitted patients | 118 | 5 | 61 | 0 | 3 | 47 | 2 |
| Patients on ≥1 antimicrobial | 98 (83.1) | 5 (100.0) | 44 (72.1) | 0 (0.0) | 3 (100.0) | 44 (93.6) | 2 (100.0) |
| **By population** |  |  |  |  |  |  |  |
| **Adult ward** |  |  |  |  |  |  |  |
| Adult medical ward |  |  |  |  |  |  |  |
| Active bed | 617 | 43 | 199 | 128 | 14 | 210 | 23 |
| Admitted patients | 436 (70.7) | 31 (72.1) | 125 (62.8) | 97 (75.8) | 13 (92.9) | 160 (76.2) | 10 (43.5) |
| Patients on ≥1 antimicrobial | 224 (51.4) | 23 (74.2) | 79 (63.2) | 40 (41.2) | 10 (76.9) | 65 (40.6) | 7 (70.0) |
| Adult surgical ward |  |  |  |  |  |  |  |
| Active bed | 300 | 0 | 145 | 39 | 0 | 110 | 6 |
| Admitted patients | 235 (78.3) | 0 (0.0) | 108 (74.5) | 32 (82.1) | 0 (0.0) | 94 (85.5) | 1 (16.7) |
| Patients on ≥1 antimicrobial | 155 (66.0) | 0 (0.0) | 80 (74.1) | 24 (75.0) | 0 (0.0) | 51 (54.3) | 0 (0.0) |
| Adult mixed medical-surgical ward |  |  |  |  |  |  |  |
| Active bed | 639 | 15 | 202 | 65 | 78 | 196 | 83 |
| Admitted patients | 446 (69.8) | 8 (53.3) | 167 (82.7) | 41 (63.1) | 41 (52.6) | 151 (77.0) | 38 (45.8) |
| Patients on ≥1 antimicrobial | 290 (65.0) | 5 (62.5) | 109 (65.3) | 28 (68.3) | 33 (80.5) | 91 (60.3) | 24 (63.2) |
| Adult ICU |  |  |  |  |  |  |  |
| Active bed | 101 | 7 | 50 | 0 | 0 | 37 | 7 |
| Admitted patients | 78 (77.2) | 3 (42.9) | 47 (94.0) | 0 (0.0) | 0 (0.0) | 26 (70.3) | 2 (28.6) |
| Patients on ≥1 antimicrobial | 58 (74.4) | 3 (100.0) | 30 (63.8) | 0 (0.0) | 0 (0.0) | 23 (88.5) | 2 (100.0) |
| **Paediatric ward** |  |  |  |  |  |  |  |
| Paediatric medical ward |  |  |  |  |  |  |  |
| Active bed | 160 | 33 | 50 | 5 | 5 | 67 | 0 |
| Admitted patients | 108 (67.5) | 12 (36.4) | 42 (84.0) | 2 (40.0) | 2 (40.0) | 50 (74.6) | 0 (0.0) |
| Patients on ≥1 antimicrobial | 67 (62.0) | 11 (91.7) | 23 (54.8) | 2 (100.0) | 1 (50.0) | 30 (60.0) | 0 (0.0) |
| Paediatric surgical ward |  |  |  |  |  |  |  |
| Active bed | 60 | 0 | 23 | 0 | 0 | 37 | 0 |
| Admitted patients | 36 (60;0) | 0 (0.0) | 11 (47.8) | 0 (0.0) | 0 (0.0) | 25 (67.6) | 0 (0.0) |
| Patients on ≥1 antimicrobial | 21 (58.3) | 0 (0.0) | 7 (63.6) | 0 (0.0) | 0 (0.0) | 14 (56.0) | 0 (0.0) |
| Paediatric mixed medical-surgical ward |  |  |  |  |  |  |  |
| Active bed | 127 | 0 | 41 | 38 | 17 | 31 | 0 |
| Admitted patients | 72 (56.7) | 0 (0.0) | 35 (85.4) | 12 (31.6) | 3 (17.60 | 22 (71.0) | 0 (0.0) |
| Patients on ≥1 antimicrobial | 41 (56.9) | 0 (0.0) | 14 (40.0) | 6 (50.0) | 2 (66.7) | 19 (86.4) | 0 (0.0) |
| Paediatric ICU |  |  |  |  |  |  |  |
| Active bed | 21 | 0 | 10 | 0 | 0 | 11 | 0 |
| Admitted patients | 19 (90.5) | 0 (0.0) | 10 (100.0) | 0 (0.0) | 0 (0.0) | 9 (81.8) | 0 (0.0) |
| Patients on ≥1 antimicrobial | 19 (100.0) | 0 (0.0) | 10 (100.0) | 0 (0.0) | 0 (0.0) | 9 (100.0) | 0 (0.0) |
| **Neonatal ward** |  |  |  |  |  |  |  |
| Neonatal ICU |  |  |  |  |  |  |  |
| Active bed | 18 | 3 | 5 | 0 | 0 | 10 | 0 |
| Admitted patients | 15 (83.3) | 2 (66.7) | 4 (80.0) | 0 (0.0) | 0 (0.0) | 9 (90.0) | 0 (0.0) |
| Patients on ≥1 antimicrobial | 15 (100.0) | 2 (100.0) | 4 (100.0) | 0 (0.0) | 0 (0.0) | 9 (100.0) | 0 (0.0) |
| **Mixed adult-neonatal-paediatric ward** |  |  |  |  |  |  |  |
| A/N/P medical ward |  |  |  |  |  |  |  |
| Active bed | 43 | 18 | 0 | 13 | 12 | 0 | 0 |
| Admitted patients | 25 (58.1) | 14 (77.8) | 0 (0.0) | 7 (53.8) | 4 (33.3) | 0 (0.0) | 0 (0.0) |
| Patients on ≥1 antimicrobial | 19 (76.0) | 13 (92.9) | 0 (0.0) | 3 (42.9) | 3 (75.0) | 0 (0.0) | 0 (0.0) |
| A/N/P surgical ward |  |  |  |  |  |  |  |
| Active bed | 41 | 0 | 0 | 0 | 0 | 41 | 0 |
| Admitted patients | 22 (53.7) | 0 (0.0) | 0 (0.0) | 0 (0.0) | 0 (0.0) | 22 (53.7) | 0 (0.0) |
| Patients on ≥1 antimicrobial | 8 (36.4) | 0 (0.0) | 0 (0.0) | 0 (0.0) | 0 (0.0) | 8 (36.4) | 0 (0.0) |
| A/N/P mixed medical-surgical ward |  |  |  |  |  |  |  |
| Active bed | 150 | 36 | 17 | 8 | 0 | 89 | 0 |
| Admitted patients | 104 (69.3) | 30 (83.3) | 13 (76.5) | 7 (87.5) | 0 (0.0) | 54 (60.7) | 0 (0.0) |
| Patients on ≥1 antimicrobial | 70 (67.3) | 18 (60.0) | 12 (92.3) | 3 (42.9) | 0 (0.0) | 37 (68.5) | 0 (0.0) |
| A/N/P ICU |  |  |  |  |  |  |  |
| Active bed | 11 | 0 | 0 | 0 | 4 | 7 | 0 |
| Admitted patients | 6 (54.5) | 0 (0.0) | 0 (0.0) | 0 (0.0) | 3 (75.0) | 3 (42.9) | 0 (0.0) |
| Patients on ≥1 antimicrobial | 6 (100.0) | 0 (0.0) | 0 (0.0) | 0 (0.0) | 3 (100.0) | 3 (100.0) | 0 (0.0) |

Abbreviations: ICU, intensive care unit; A/N/P, adult-neonatal-paediatric

This table includes antimicrobials for systemic use (J01, J02, J02, J04, J05, P01AB, A07A, P1B).

**Table S2. Systemic antibiotic use by hospital**

| **Systemic antibiotic (J01)** | **Total**  **(n=1,273)** | **Hospital 1**  **(n=91)** | **Hospital 2**  **(n=517)** | **Hospital 3**  **(n=126)** | **Hospital 4**  **(n=62)** | **Hospital 5**  **(n=446)** | **Hospital 6**  **(n=31)** |
| --- | --- | --- | --- | --- | --- | --- | --- |
| Ceftriaxone | 341 (26.8) | 41 (45.1) | 178 (34.4) | 29 (23.0) | 22 (35.5) | 60 (13.5) | 11 (35.5) |
| Levofloxacin | 136 (10.7) | 12 (13.2) | 67 (13.0) | 16 (12.7) | 0 (0.0) | 39 (8.7) | 2 (6.5) |
| Metronidazole | 90 (7.1) | 1 (1.1) | 43 (8.3) | 6 (4.8) | 6 (9.7) | 32 (7.2) | 2 (6.5) |
| Meropenem | 82 (6.4) | 6 (6.6) | 10 (1.9) | 20 (15.9) | 4 (6.5) | 33 (7.4) | 9 (29.0) |
| Cefotaxime | 71 (5.6) | 2 (2.2) | 36 (7.0) | 3 (2.4) | 3 (4.8) | 27 (6.1) | 0 (0.0) |
| Cefoperazone | 52 (4.1) | 0 (0.0) | 28 (5.4) | 1 (0.8) | 1 (1.6) | 22 (4.9) | 0 (0.0) |
| Cefixime | 51 (4.0) | 6 (6.6) | 27 (5.2) | 11 (8.7) | 0 (0.0) | 6 (1.3) | 1 (3.2) |
| Ampicillin sulbactam | 44 (3.5) | 0 (0.0) | 5 (1.0) | 1 (0.8) | 1 (1.6) | 37 (8.3) | 0 (0.0) |
| Gentamicin | 42 (3.3) | 0 (0.0) | 21 (4.1) | 1 (0.8) | 2 (3.2) | 18 (4.0) | 0 (0.0) |
| Amikacin | 32 (2.5) | 1 (1.1) | 13 (2.5) | 3 (2.4) | 0 (0.0) | 15 (3.4) | 0 (0.0) |
| Amoxicillin + clavulanic acid | 31 (2.4) | 0 (0.0) | 23 (4.4) | 0 (0.0) | 1 (1.6) | 7 (1.6) | 0 (0.0) |
| Cefoperazone sulbactam | 28 (2.2) | 2 (2.2) | 0 (0.0) | 0 (0.0) | 0 (0.0) | 25 (5.6) | 1 (3.2) |
| Co-trimoxazole | 28 (2.2) | 3 (3.3) | 4 (0.8) | 3 (2.4) | 3 (4.8) | 14 (3.1) | 1 (3.2) |
| Ceftazidime | 21 (1.6) | 0 (0.0) | 3 (0.6) | 4 (3.2) | 1 (1.6) | 13 (2.9) | 0 (0.0) |
| Ciprofloxacin | 21 (1.6) | 1 (1.1) | 8 (1.5) | 2 (1.6) | 0 (0.0) | 8 (1.8) | 2 (6.5) |
| Cefazolin | 18 (1.4) | 0 (0.0) | 9 (1.7) | 2 (1.6) | 0 (0.0) | 7 (1.6) | 0 (0.0) |
| Ampicillin | 17 (1.3) | 0 (0.0) | 2 (0.4) | 0 (0.0) | 5 (8.1) | 10 (2.2) | 0 (0.0) |
| Amoxicillin | 16 (1.3) | 0 (0.0) | 13 (2.5) | 2 (1.6) | 0 (0.0) | 1 (0.2) | 0 (0.0) |
| Azithromycin | 16 (1.3) | 0 (0.0) | 4 (0.8) | 3 (2.4) | 1 (1.6) | 8 (1.8) | 0 (0.0) |
| Clindamycin | 16 (1.3) | 1 (1.1) | 3 (0.6) | 3 (2.4) | 4 (6.5) | 5 (1.1) | 0 (0.0) |
| Fosfomycin | 15 (1.2) | 0 (0.0) | 5 (1.0) | 0 (0.0) | 0 (0.0) | 10 (2.2) | 0 (0.0) |
| Moxifloxacin | 13 (1.0) | 2 (2.2) | 2 (0.4) | 7 (5.6) | 1 (1.6) | 1 (0.2) | 0 (0.0) |
| Tigecycline | 13 (1.0) | 0 (0.0) | 0 (0.0) | 3 (2.4) | 0 (0.0) | 10 (2.2) | 0 (0.0) |
| Streptomycin | 10 (0.8) | 0 (0.0) | 2 (0.4) | 0 (0.0) | 3 (4.8) | 5 (1.1) | 0 (0.0) |
| Cefadroxil | 9 (0.7) | 3 (3.3) | 3 (0.6) | 0 (0.0) | 0 (0.0) | 3 (0.7) | 0 (0.0) |
| Imipenem and cilastatin | 7 (0.5) | 2 (2.2) | 1 (0.2) | 3 (2.4) | 0 (0.0) | 1 (0.2) | 0 (0.0) |
| Piperacilin tazobactam | 7 (0.5) | 0 (0.0) | 0 (0.0) | 0 (0.0) | 0 (0.0) | 7 (1.6) | 0 (0.0) |
| Cefepime | 6 (0.5) | 1 (1.1) | 1 (0.2) | 0 (0.0) | 1 (1.6) | 3 (0.7) | 0 (0.0) |
| Doripenem | 5 (0.4) | 0 (0.0) | 0 (0.0) | 0 (0.0) | 0 (0.0) | 4 (0.9) | 1 (3.2) |
| Erythromycin | 5 (0.4) | 2 (2.2) | 0 (0.0) | 0 (0.0) | 1 (1.6) | 2 (0.4) | 0 (0.0) |
| Sultamicillin | 5 (0.4) | 2 (2.2) | 1 (0.2) | 0 (0.0) | 0 (0.0) | 2 (0.4) | 0 (0.0) |
| Clarithromycin | 4 (0.3) | 0 (0.0) | 0 (0.0) | 0 (0.0) | 0 (0.0) | 3 (0.7) | 1 (3.2) |
| Cefuroxime | 3 (0.2) | 0 (0.0) | 0 (0.0) | 3 (2.4) | 0 (0.0) | 0 (0.0) | 0 (0.0) |
| Chloramphenicol | 3 (0.2) | 0 (0.0) | 2 (0.4) | 0 (0.0) | 0 (0.0) | 1 (0.2) | 0 (0.0) |
| Vancomycin | 3 (0.2) | 0 (0.0) | 1 (0.2) | 0 (0.0) | 0 (0.0) | 2 (0.4) | 0 (0.0) |
| Penicillin procaine | 2 (0.2) | 0 (0.0) | 0 (0.0) | 0 (0.0) | 2 (3.2) | 0 (0.0) | 0 (0.0) |
| Cefditoren pivoxil | 1 (0.1) | 1 (1.1) | 0 (0.0) | 0 (0.0) | 0 (0.0) | 0 (0.0) | 0 (0.0) |
| Colistin | 1 (0.1) | 0 (0.0) | 1 (0.2) | 0 (0.0) | 0 (0.0) | 0 (0.0) | 0 (0.0) |
| Doxycycline | 1 (0.1) | 0 (0.0) | 0 (0.0) | 0 (0.0) | 0 (0.0) | 1 (0.2) | 0 (0.0) |
| Gentamicin | 1 (0.1) | 0 (0.0) | 0 (0.0) | 0 (0.0) | 0 (0.0) | 1 (0.2) | 0 (0.0) |
| Levofloxacin | 1 (0.1) | 0 (0.0) | 0 (0.0) | 0 (0.0) | 0 (0.0) | 1 (0.2) | 0 (0.0) |
| Linezolid | 1 (0.1) | 1 (1.1) | 0 (0.0) | 0 (0.0) | 0 (0.0) | 0 (0.0) | 0 (0.0) |
| Metronidazole | 1 (0.1) | 0 (0.0) | 0 (0.0) | 0 (0.0) | 0 (0.0) | 1 (0.2) | 0 (0.0) |
| Netilmicin | 1 (0.1) | 0 (0.0) | 1 (0.2) | 0 (0.0) | 0 (0.0) | 0 (0.0) | 0 (0.0) |
| Ofloxacin | 1 (0.1) | 1 (1.1) | 0 (0.0) | 0 (0.0) | 0 (0.0) | 0 (0.0) | 0 (0.0) |
| Polymyxin B | 1 (0.1) | 0 (0.0) | 0 (0.0) | 0 (0.0) | 0 (0.0) | 1 (0.2) | 0 (0.0) |

**Table S3. Diagnosis/reason for systemic antimicrobial use by hospital**

| **Diagnosis** | **Total (n=1,666)** | **Hospital 1**  **(n=114)** | **Hospital 2**  **(n=630)** | **Hospital 3**  **(n=158)** | **Hospital 4**  **(n=98)** | **Hospital 5**  **(n=622)** | **Hospital 6**  **(n=44)** |
| --- | --- | --- | --- | --- | --- | --- | --- |
| Pneumonia or lower respiratory tract infection | 365 (21.9) | 16 (14.0) | 155 (24.6) | 42 (26.6) | 17 (17.3) | 129 (20.7) | 6 (13.6) |
| Pulmonary tuberculosis | 185 (11.1) | 8 (7.0) | 66 (10.5) | 13 (8.2) | 26 (26.5) | 66 (10.6) | 6 (13.6) |
| Skin and soft tissue infection ^a^ | 110 (6.6) | 4 (3.5) | 45 (7.1) | 10 (6.3) | 7 (7.1) | 42 (6.8) | 2 (4.5) |
| Prophylaxis for gastrointestinal infection ^b^ | 102 (6.1) | 1 (0.9) | 52 (8.3) | 10 (6.3) | 1 (1.0) | 35 (5.6) | 3 (6.8) |
| Gastrointestinal infection | 84 (5.0) | 26 (22.8) | 13 (2.1) | 10 (6.3) | 1 (1.0) | 27 (4.3) | 7 (15.9) |
| Unknown reason | 79 (4.7) | 2 (1.8) | 30 (4.8) | 6 (3.8) | 8 (8.2) | 29 (4.7) | 4 (9.1) |
| Central nervous system infection | 69 (4.1) | 0 (0.0) | 23 (3.7) | 3 (1.9) | 9 (9.2) | 30 (4.8) | 4 (9.1) |
| Intra-abdominal infection ^c^ | 66 (4.0) | 0 (0.0) | 36 (5.7) | 2 (1.3) | 0 (0.0) | 27 (4.3) | 1 (2.3) |
| Prophylaxis for bone and joint infection ^d^ | 60 (3.6) | 2 (1.8) | 27 (4.3) | 8 (5.1) | 2 (2.0) | 20 (3.2) | 1 (2.3) |
| Prophylaxis for obstetric or gynaecological infection | 58 (3.5) | 1 (0.9) | 39 (6.2) | 9 (5.7) | 3 (3.1) | 6 (1.0) | 0 (0.0) |
| Human Immunodeficiency Virus | 49 (2.9) | 6 (5.3) | 8 (1.3) | 4 (2.5) | 8 (8.2) | 23 (3.7) | 0 (0.0) |
| Sepsis | 49 (2.9) | 4 (3.5) | 23 (3.7) | 4 (2.5) | 0 (0.0) | 18 (2.9) | 0 (0.0) |
| Prophylaxis for urinary tract infection (surgery or recurrent infection) | 42 (2.5) | 1 (0.9) | 14 (2.2) | 5 (3.2) | 0 (0.0) | 21 (3.4) | 1 (2.3) |
| General medical prophylaxis | 41 (2.5) | 6 (5.3) | 12 (1.9) | 3 (1.9) | 4 (4.1) | 14 (2.3) | 2 (4.5) |
| Ear, nose, throat infection ^e^ | 40 (2.4) | 5 (4.4) | 9 (1.4) | 4 (2.5) | 4 (4.1) | 18 (2.9) | 0 (0.0) |
| Other diagnosis ^f^ | 36 (2.2) | 11 (9.6) | 12 (1.9) | 5 (3.2) | 0 (0.0) | 7 (1.1) | 1 (2.3) |
| Upper urinary tract infection ^g^ | 35 (2.1) | 0 (0.0) | 11 (1.7) | 3 (1.9) | 0 (0.0) | 21 (3.4) | 0 (0.0) |
| Medical prophylaxis for new-born risk factors | 28 (1.7) | 0 (0.0) | 8 (1.3) | 0 (0.0) | 2 (2.0) | 18 (2.9) | 0 (0.0) |
| Prophylaxis for ear, nose, throat infection | 26 (1.6) | 3 (2.6) | 12 (1.9) | 1 (0.6) | 0 (0.0) | 7 (1.1) | 3 (6.8) |
| Prophylaxis for central nervous system infection | 25 (1.5) | 3 (2.6) | 13 (2.1) | 0 (0.0) | 0 (0.0) | 9 (1.4) | 0 (0.0) |
| Prophylaxis for respiratory infection | 22 (1.3) | 2 (1.8) | 5 (0.8) | 3 (1.9) | 4 (4.1) | 8 (1.3) | 0 (0.0) |
| Lower urinary tract infections (cystitis) | 20 (1.2) | 4 (3.5) | 3 (0.5) | 7 (4.4) | 0 (0.0) | 5 (0.8) | 1 (2.3) |
| Bone or joint infection ^h^ | 13 (0.8) | 1 (0.9) | 5 (0.8) | 0 (0.0) | 0 (0.0) | 7 (1.1) | 0 (0.0) |
| Febrile neutropenia | 11 (0.7) | 1 (0.9) | 0 (0.0) | 0 (0.0) | 0 (0.0) | 10 (1.6) | 0 (0.0) |
| Upper respiratory tract viral infection ^i^ | 9 (0.5) | 7 (6.1) | 0 (0.0) | 1 (0.6) | 0 (0.0) | 0 (0.0) | 1 (2.3) |
| Prophylaxis for cardiac or vascular infection | 8 (0.5) | 0 (0.0) | 0 (0.0) | 0 (0.0) | 0 (0.0) | 8 (1.3) | 0 (0.0) |
| Cardiovascular system infections | 7 (0.4) | 0 (0.0) | 2 (0.3) | 0 (0.0) | 0 (0.0) | 5 (0.8) | 0 (0.0) |
| Prophylaxis for eye operations | 5 (0.3) | 0 (0.0) | 1 (0.2) | 0 (0.0) | 0 (0.0) | 4 (0.6) | 0 (0.0) |
| Acute bronchitis or exacerbations of chronic bronchitis | 4 (0.2) | 0 (0.0) | 3 (0.5) | 0 (0.0) | 1 (1.0) | 0 (0.0) | 0 (0.0) |
| Medical prophylaxis for maternal risk factors | 4 (0.2) | 0 (0.0) | 0 (0.0) | 1 (0.6) | 0 (0.0) | 3 (0.5) | 0 (0.0) |
| Bacteraemia | 3 (0.2) | 0 (0.0) | 0 (0.0) | 0 (0.0) | 0 (0.0) | 2 (0.3) | 1 (2.3) |
| Pyrexia of unknown origin | 3 (0.2) | 0 (0.0) | 1 (0.2) | 1 (0.6) | 1 (1.0) | 0 (0.0) | 0 (0.0) |
| Malaria | 2 (0.1) | 0 (0.0) | 0 (0.0) | 2 (1.3) | 0 (0.0) | 0 (0.0) | 0 (0.0) |
| Obstetric/gynaecological infections ^j^ | 2 (0.1) | 0 (0.0) | 1 (0.2) | 0 (0.0) | 0 (0.0) | 1 (0.2) | 0 (0.0) |
| Pyrexia of unknown origin in non-neutropenic haemato-oncology patients | 2 (0.1) | 0 (0.0) | 1 (0.2) | 0 (0.0) | 0 (0.0) | 1 (0.2) | 0 (0.0) |
| Eye infections | 1 (0.1) | 0 (0.0) | 0 (0.0) | 1 (0.6) | 0 (0.0) | 0 (0.0) | 0 (0.0) |
| Genito-urinary infections ^k^ | 1 (0.1) | 0 (0.0) | 0 (0.0) | 0 (0.0) | 0 (0.0) | 1 (0.2) | 0 (0.0) |

The table lists the most common reasons to prescribe at least one antibiotic for systemic use (J01, J02, J02, J04, J05, P01AB, A07A, P01B). Data are expressed as numbers (percentage) and ranked by frequency. Patients recorded with more than one diagnosis were counted by number of diagnoses. Diagnosis were coded based on the GLOBAL-PPS 2018 Diagnostic Code List.

^a^	Including cellulitis, wound including surgical site infections, deep soft tissue not involving bone (e.g. infected pressure or diabetic ulcers, abscess).

^b^	Including prophylaxis for surgery of the gastrointestinal tract, liver, or biliary tree, and prophylaxis in patients with neutropenia or hepatic failure.

^c^	Including hepatobiliary, intra-abdominal abscess, etc.

^d^	Including prophylaxis for surgical site infections, for plastic or orthopaedic surgery (bone or joint).

^e^	Including mouth, sinuses, larynx.

^f.^	Antibiotic prescribed with documentation for which there is no above diagnosis group.

^g^	Including catheter related urinary tract infection, pyelonephritis.

^h^	Including septic arthritis, including prosthetic joint, osteomyelitis.

^i^	Including influenza but not ear, nose, throat infections.

^j^	Including sexually transmitted infections in women.

^k^	Including sexually transmitted infections in men.

**Table S4. Diagnosis/reason for systemic antibiotic use by indication**

| **Diagnosis** | **Total**  **(n=1,273)** | **CAI**  **(n=542)** | **HAI**  **(n=235)** | **Medical**  **prophylaxis**  **(n=122)** | **Surgical prophylaxis**  **(n=288)** | **Other indication ^l^**  **(n=27)** | **Unknown**  **Indication**  **(n=59)** |
| --- | --- | --- | --- | --- | --- | --- | --- |
| Pneumonia or lower respiratory tract infection | 353 (27.7) | 231 (42.6) | 122 (51.9) | 0 (0.0) | 0 (0.0) | 0 (0.0) | 0 (0.0) |
| Skin and soft tissue infection ^a^ | 106 (8.3) | 77 (14.2) | 29 (12.3) | 0 (0.0) | 0 (0.0) | 0 (0.0) | 0 (0.0) |
| Prophylaxis for gastrointestinal infections ^b^ | 101 (7.9) | 0 (0.0) | 0 (0.0) | 21 (17.2) | 80 (27.8) | 0 (0.0) | 0 (0.0) |
| Gastrointestinal infection | 69 (5.4) | 66 (12.2) | 3 (1.3) | 0 (0.0) | 0 (0.0) | 0 (0.0) | 0 (0.0) |
| Prophylaxis for bone and joint infection ^c^ | 58 (4.6) | 0 (0.0) | 0 (0.0) | 9 (7.4) | 49 (17.0) | 0 (0.0) | 0 (0.0) |
| Prophylaxis for obstetrics or gynaecological infection | 58 (4.6) | 0 (0.0) | 0 (0.0) | 0 (0.0) | 58 (20.1) | 0 (0.0) | 0 (0.0) |
| Intra-abdominal infection ^d^ | 57 (4.5) | 29 (5.4) | 28 (11.9) | 0 (0.0) | 0 (0.0) | 0 (0.0) | 0 (0.0) |
| Unknown reason | 47 (3.7) | 0 (0.0) | 0 (0.0) | 10 (8.2) | 0 (0.0) | 0 (0.0) | 37 (62.7) |
| Sepsis | 46 (3.6) | 30 (5.5) | 16 (6.8) | 0 (0.0) | 0 (0.0) | 0 (0.0) | 0 (0.0) |
| Prophylaxis for urinary tract infection (surgery or recurrent infection) | 42 (3.3) | 0 (0.0) | 0 (0.0) | 5 (4.1) | 37 (12.8) | 0 (0.0) | 0 (0.0) |
| Other diagnosis ^e^ | 35 (2.7) | 4 (0.7) | 0 (0.0) | 1 (0.8) | 8 (2.8) | 0 (0.0) | 22 (37.3) |
| Ear, nose, throat infection ^f^ | 30 (2.4) | 28 (5.2) | 2 (0.9) | 0 (0.0) | 0 (0.0) | 0 (0.0) | 0 (0.0) |
| Upper urinary tract infection ^g^ | 29 (2.3) | 8 (1.5) | 21 (8.9) | 0 (0.0) | 0 (0.0) | 0 (0.0) | 0 (0.0) |
| Central nervous system infection | 26 (2.0) | 11 (2.0) | 2 (0.9) | 0 (0.0) | 0 (0.0) | 13 (48.1) | 0 (0.0) |
| Medical prophylaxis for new-born risk factors | 25 (2.0) | 0 (0.0) | 0 (0.0) | 25 (20.5) | 0 (0.0) | 0 (0.0) | 0 (0.0) |
| General medical prophylaxis | 24 (1.9) | 0 (0.0) | 0 (0.0) | 24 (19.7) | 0 (0.0) | 0 (0.0) | 0 (0.0) |
| Prophylaxis central nervous system infection | 24 (1.9) | 0 (0.0) | 0 (0.0) | 3 (2.5) | 21 (7.3) | 0 (0.0) | 0 (0.0) |
| Prophylaxis ear, nose, throat infection | 22 (1.7) | 0 (0.0) | 0 (0.0) | 1 (0.8) | 21 (7.3) | 0 (0.0) | 0 (0.0) |
| Prophylaxis for respiratory infection | 20 (1.6) | 0 (0.0) | 0 (0.0) | 19 (15.6) | 1 (0.3) | 0 (0.0) | 0 (0.0) |
| Lower urinary tract infections (cystitis) | 19 (1.5) | 17 (3.1) | 2 (0.9) | 0 (0.0) | 0 (0.0) | 0 (0.0) | 0 (0.0) |
| Pulmonary tuberculosis | 14 (1.1) | 0 (0.0) | 0 (0.0) | 0 (0.0) | 0 (0.0) | 14 (51.9) | 0 (0.0) |
| Bone and joint infection ^h^ | 13 (1.0) | 12 (2.2) | 1 (0.4) | 0 (0.0) | 0 (0.0) | 0 (0.0) | 0 (0.0) |
| Upper respiratory tract viral infection ^i^ | 9 (0.7) | 9 (1.7) | 0 (0.0) | 0 (0.0) | 0 (0.0) | 0 (0.0) | 0 (0.0) |
| Prophylaxis for cardiac or vascular infection | 8 (0.6) | 0 (0.0) | 0 (0.0) | 0 (0.0) | 8 (2.8) | 0 (0.0) | 0 (0.0) |
| Cardiovascular system infection | 7 (0.5) | 7 (1.3) | 0 (0.0) | 0 (0.0) | 0 (0.0) | 0 (0.0) | 0 (0.0) |
| Febrile neutropenia | 6 (0.5) | 4 (0.7) | 2 (0.9) | 0 (0.0) | 0 (0.0) | 0 (0.0) | 0 (0.0) |
| Prophylaxis for eye operation | 5 (0.4) | 0 (0.0) | 0 (0.0) | 0 (0.0) | 5 (1.7) | 0 (0.0) | 0 (0.0) |
| Acute bronchitis or exacerbations of chronic bronchitis | 4 (0.3) | 1 (0.2) | 3 (1.3) | 0 (0.0) | 0 (0.0) | 0 (0.0) | 0 (0.0) |
| Medical prophylaxis for maternal risk factors | 4 (0.3) | 0 (0.0) | 0 (0.0) | 4 (3.3) | 0 (0.0) | 0 (0.0) | 0 (0.0) |
| Bacteraemia | 3 (0.2) | 1 (0.2) | 2 (0.9) | 0 (0.0) | 0 (0.0) | 0 (0.0) | 0 (0.0) |
| Pyrexia of unknown origin | 3 (0.2) | 2 (0.4) | 1 (0.4) | 0 (0.0) | 0 (0.0) | 0 (0.0) | 0 (0.0) |
| Obstetrics/Gynaecological infections ^j^ | 2 (0.2) | 2 (0.4) | 0 (0.0) | 0 (0.0) | 0 (0.0) | 0 (0.0) | 0 (0.0) |
| Pyrexia of unknown origin in non-neutropenic haemato-oncology patients | 2 (0.2) | 1 (0.2) | 1 (0.4) | 0 (0.0) | 0 (0.0) | 0 (0.0) | 0 (0.0) |
| Eye infections | 1 (0.1) | 1 (0.2) | 0 (0.0) | 0 (0.0) | 0 (0.0) | 0 (0.0) | 0 (0.0) |
| Genito-urinary infections ^k^ | 1 (0.1) | 1 (0.2) | 0 (0.0) | 0 (0.0) | 0 (0.0) | 0 (0.0) | 0 (0.0) |

The table lists the most common reasons to prescribe at least one antibiotic for systemic use (J01). Data are expressed as numbers (percentage) and ranked by frequency. Patients recorded with more than one diagnosis were counted by number of diagnoses. Diagnosis were coded based on the GLOBAL-PPS 2018 Diagnostic Code List.

^a^ Including cellulitis, wound including surgical site infections, deep soft tissue not involving bone (e.g. infected pressure or diabetic ulcers, abscess).

^b^ Including prophylaxis for surgery of the gastrointestinal tract, liver, or biliary tree, and prophylaxis in patients with neutropenia or hepatic failure.

^c^ Including prophylaxis for surgical site infections, for plastic or orthopaedic surgery (bone or joint).

^d^ Including hepatobiliary, intra-abdominal abscess, etc.

^e^ Antibiotic prescribed with documentation for which there is no above diagnosis group.

^f^ Including mouth, sinuses, larynx.

^g^ Including catheter related urinary tract infection, pyelonephritis.

^h^ Including septic arthritis, including prosthetic joint, osteomyelitis.

^I^ Including influenza but not ear, nose, throat infections.

^j^ Including sexually transmitted infections in women.

^k^ Including sexually transmitted infections in men.

^l^ Other indication included antibiotics prescribed for neurotoxoplasmosis, pulmonary tuberculosis, and as motility agent.

Abbreviations: CAI, community acquired infection; HAI, hospital acquired infection;

**Table S5. Systemic antibiotic use (J01) by indication**

| **Hospital** | **Total**  **(n = 1,273)** | **CAI Empirical**  **prescribing**  **(n = 498)** | **CAI**  **Targeted**  **prescribing**  **(n = 44)** | **HAI**  **Empirical**  **prescribing**  **(n = 172)** | **HAI**  **Targeted**  **prescribing**  **(n = 63)** | **Medical**  **prophylaxis**  **(n = 122)** | **Surgical prophylaxis**  **(n = 288)** | **Other indication^a^**  **(n = 27)** | **Unknown**  **indication**  **(n = 59)** |
| --- | --- | --- | --- | --- | --- | --- | --- | --- | --- |
| Hospital 1 | 91 (7.1) | 59 (11.8) | 2 (4.5) | 3 (1.7) | 1 (1.6) | 9 (7.4) | 4 (1.4) | 1 (3.7) | 12 (20.3) |
| Hospital 2 | 517 (40.6) | 199 (40.0) | 12 (27.3) | 62 (36.0) | 20 (31.7) | 50 (41.0) | 146 (50.7) | 6 (22.2) | 22 (37.3) |
| Hospital 3 | 126 (9.9) | 61 (12.2) | 4 (9.1) | 14 (8.1) | 6 (9.5) | 4 (3.3) | 31 (10.8) | 1 (3.7) | 5 (8.5) |
| Hospital 4 | 62 (4.9) | 27 (5.4) | 1 (2.3) | 2 (1.2) | 1 (1.6) | 7 (5.7) | 7 (2.4) | 9 (33.3) | 8 (13.6) |
| Hospital 5 | 446 (35.0) | 139 (27.9) | 23 (52.3) | 90 (52.3) | 32 (50.8) | 50 (41.0) | 93 (32.3) | 10 (37.0) | 9 (15.3) |
| Hospital 6 | 31 (2.4) | 13 (2.6) | 2 (4.5) | 1 (0.6) | 3 (4.8) | 2 (1.6) | 7 (2.4) | 0 (0.0) | 3 (5.1) |

This table includes antibiotics for systemic use (J01).

Abbreviations: CAI, community acquired infection; HAI, hospital acquired infection;

^a^ Other indication included antibiotics prescribed for neurotoxoplasmosis, pulmonary tuberculosis, and as motility agent.

**Table S6. Systemic antibiotic use, by indication**

| **Systemic antibiotic (J01)** | **Total**  **(n=1,273)** | **CAI**  **(n=542)** | **HAI**  **(n=235)** | **Medical**  **prophylaxis**  **(n=122)** | **Surgical prophylaxis**  **(n=288)** | **Other indication ^a^**  **(n=27)** | **Unknown**  **Indication**  **(n=59)** |
| --- | --- | --- | --- | --- | --- | --- | --- |
| Ceftriaxone | 341 (26.8) | 178 (32.8) | 23 (9.8) | 35 (28.7) | 76 (26.4) | 0 (0.0) | 29 (49.2) |
| Levofloxacin | 136 (10.7) | 73 (13.5) | 44 (18.7) | 3 (2.5) | 8 (2.8) | 3 (11.1) | 5 (8.5) |
| Metronidazole | 90 (7.1) | 44 (8.1) | 14 (6.0) | 2 (1.6) | 28 (9.7) | 0 (0.0) | 2 (3.4) |
| Meropenem | 82 (6.4) | 42 (7.7) | 32 (13.6) | 2 (1.6) | 3 (1.0) | 1 (3.7) | 2 (3.4) |
| Cefotaxime | 71 (5.6) | 27 (5.0) | 12 (5.1) | 10 (8.2) | 14 (4.9) | 1 (3.7) | 7 (11.9) |
| Cefoperazone | 52 (4.1) | 11 (2.0) | 5 (2.1) | 4 (3.3) | 32 (11.1) | 0 (0.0) | 0 (0.0) |
| Cefixime | 51 (4.0) | 12 (2.2) | 3 (1.3) | 2 (1.6) | 33 (11.5) | 0 (0.0) | 1 (1.7) |
| Ampicillin sulbactam | 44 (3.5) | 29 (5.4) | 5 (2.1) | 4 (3.3) | 5 (1.7) | 0 (0.0) | 1 (1.7) |
| Gentamicin | 42 (3.3) | 17 (3.1) | 3 (1.3) | 13 (10.7) | 7 (2.4) | 0 (0.0) | 2 (3.4) |
| Amikacin | 32 (2.5) | 10 (1.8) | 15 (6.4) | 0 (0.0) | 6 (2.1) | 0 (0.0) | 1 (1.7) |
| Amoxicillin + clavulanic acid | 31 (2.4) | 8 (1.5) | 5 (2.1) | 1 (0.8) | 16 (5.6) | 0 (0.0) | 1 (1.7) |
| Cefoperazone sulbactam | 28 (2.2) | 9 (1.7) | 6 (2.6) | 6 (4.9) | 6 (2.1) | 0 (0.0) | 1 (1.7) |
| Co-trimoxazole | 28 (2.2) | 4 (0.7) | 2 (0.9) | 21 (17.2) | 1 (0.3) | 0 (0.0) | 0 (0.0) |
| Ceftazidime | 21 (1.6) | 5 (0.9) | 14 (6.0) | 0 (0.0) | 2 (0.7) | 0 (0.0) | 0 (0.0) |
| Ciprofloxacin | 21 (1.6) | 9 (1.7) | 9 (3.8) | 1 (0.8) | 2 (0.7) | 0 (0.0) | 0 (0.0) |
| Cefazolin | 18 (1.4) | 0 (0.0) | 0 (0.0) | 0 (0.0) | 18 (6.2) | 0 (0.0) | 0 (0.0) |
| Ampicillin | 17 (1.3) | 3 (0.6) | 0 (0.0) | 9 (7.4) | 4 (1.4) | 0 (0.0) | 1 (1.7) |
| Amoxicillin | 16 (1.3) | 7 (1.3) | 0 (0.0) | 4 (3.3) | 3 (1.0) | 0 (0.0) | 2 (3.4) |
| Azithromycin | 16 (1.3) | 9 (1.7) | 6 (2.6) | 0 (0.0) | 0 (0.0) | 0 (0.0) | 1 (1.7) |
| Clindamycin | 16 (1.3) | 1 (0.2) | 1 (0.4) | 1 (0.8) | 3 (1.0) | 10 (37.0) | 0 (0.0) |
| Fosfomycin | 15 (1.2) | 4 (0.7) | 5 (2.1) | 0 (0.0) | 6 (2.1) | 0 (0.0) | 0 (0.0) |
| Moxifloxacin | 13 (1.0) | 8 (1.5) | 3 (1.3) | 0 (0.0) | 2 (0.7) | 0 (0.0) | 0 (0.0) |
| Tigecycline | 13 (1.0) | 7 (1.3) | 6 (2.6) | 0 (0.0) | 0 (0.0) | 0 (0.0) | 0 (0.0) |
| Streptomycin | 10 (0.8) | 0 (0.0) | 0 (0.0) | 0 (0.0) | 0 (0.0) | 10 (37.0) | 0 (0.0) |
| Cefadroxil | 9 (0.7) | 4 (0.7) | 0 (0.0) | 0 (0.0) | 4 (1.4) | 0 (0.0) | 1 (1.7) |
| Imipenem and cilastatin | 7 (0.5) | 4 (0.7) | 2 (0.9) | 0 (0.0) | 1 (0.3) | 0 (0.0) | 0 (0.0) |
| Piperacilin tazobactam | 7 (0.5) | 1 (0.2) | 5 (2.1) | 0 (0.0) | 0 (0.0) | 0 (0.0) | 1 (1.7) |
| Cefepime | 6 (0.5) | 4 (0.7) | 1 (0.4) | 0 (0.0) | 1 (0.3) | 0 (0.0) | 0 (0.0) |
| Doripenem | 5 (0.4) | 0 (0.0) | 4 (1.7) | 0 (0.0) | 0 (0.0) | 0 (0.0) | 1 (1.7) |
| Erythromycin | 5 (0.4) | 2 (0.4) | 0 (0.0) | 3 (2.5) | 0 (0.0) | 0 (0.0) | 0 (0.0) |
| Sultamicillin | 5 (0.4) | 3 (0.6) | 2 (0.9) | 0 (0.0) | 0 (0.0) | 0 (0.0) | 0 (0.0) |
| Clarithromycin | 4 (0.3) | 1 (0.2) | 2 (0.9) | 1 (0.8) | 0 (0.0) | 0 (0.0) | 0 (0.0) |
| Cefuroxime | 3 (0.2) | 0 (0.0) | 0 (0.0) | 0 (0.0) | 3 (1.0) | 0 (0.0) | 0 (0.0) |
| Chloramphenicol | 3 (0.2) | 2 (0.4) | 0 (0.0) | 0 (0.0) | 1 (0.3) | 0 (0.0) | 0 (0.0) |
| Vancomycin | 3 (0.2) | 1 (0.2) | 2 (0.9) | 0 (0.0) | 0 (0.0) | 0 (0.0) | 0 (0.0) |
| Penicillin procaine | 2 (0.2) | 2 (0.4) | 0 (0.0) | 0 (0.0) | 0 (0.0) | 0 (0.0) | 0 (0.0) |
| Cefditoren pivoxil | 1 (0.1) | 0 (0.0) | 0 (0.0) | 0 (0.0) | 1 (0.3) | 0 (0.0) | 0 (0.0) |
| Colistin | 1 (0.1) | 0 (0.0) | 1 (0.4) | 0 (0.0) | 0 (0.0) | 0 (0.0) | 0 (0.0) |
| Doxycycline | 1 (0.1) | 0 (0.0) | 0 (0.0) | 0 (0.0) | 0 (0.0) | 1 (3.7) | 0 (0.0) |
| Gentamicin | 1 (0.1) | 0 (0.0) | 1 (0.4) | 0 (0.0) | 0 (0.0) | 0 (0.0) | 0 (0.0) |
| Levofloxacin | 1 (0.1) | 0 (0.0) | 0 (0.0) | 0 (0.0) | 1 (0.3) | 0 (0.0) | 0 (0.0) |
| Linezolid | 1 (0.1) | 1 (0.2) | 0 (0.0) | 0 (0.0) | 0 (0.0) | 0 (0.0) | 0 (0.0) |
| Metronidazole | 1 (0.1) | 0 (0.0) | 1 (0.4) | 0 (0.0) | 0 (0.0) | 0 (0.0) | 0 (0.0) |
| Netilmicin | 1 (0.1) | 0 (0.0) | 0 (0.0) | 0 (0.0) | 1 (0.3) | 0 (0.0) | 0 (0.0) |
| Ofloxacin | 1 (0.1) | 0 (0.0) | 0 (0.0) | 0 (0.0) | 0 (0.0) | 1 (3.7) | 0 (0.0) |
| Polymyxin B | 1 (0.1) | 0 (0.0) | 1 (0.4) | 0 (0.0) | 0 (0.0) | 0 (0.0) | 0 (0.0) |

This table includes antibiotics for systemic use (J01).

^a^ Other indication included antibiotics prescribed for neurotoxoplasmosis, pulmonary tuberculosis, and as motility agent.

Abbreviations: CAI, community acquired infection; HAI, hospital acquired infection;

**Table S7. Systemic antibiotic use for hospital-acquired infections**

| **Diagnosis** | **Total**  **(n=235)** | **Post-operative surgical site infection ^f^ (n=32)** | **Intervention related infections ^g^ (n=35)** | **C. difficile associated diarrhoea ^h^ (n=0)** | **Other hospital acquired infection ^i^ (incl. HAP) (n=165)** | **Infection present on transfer-in from other hospital ^j^ (n=2)** | **Infection present on transfer from long-term care facility ^k^ (n=1)** |
| --- | --- | --- | --- | --- | --- | --- | --- |
| Pneumonia or lower respiratory tract infection | 122 (51.9) | 0 (0.0) | 11 (31.4) | 0 (0.0) | 108 (65.5) | 2 (100.0) | 1 (100.0) |
| Skin and soft tissue infection ^a^ | 29 (12.3) | 25 (78.1) | 0 (0.0) | 0 (0.0) | 4 (2.4) | 0 (0.0) | 0 (0.0) |
| Intra-abdominal Infection ^b^ | 28 (11.9) | 5 (15.6) | 5 (14.3) | 0 (0.0) | 18 (10.9) | 0 (0.0) | 0 (0.0) |
| Upper urinary tract infection ^c^ | 21 (8.9) | 1 (3.1) | 13 (37.1) | 0 (0.0) | 7 (4.2) | 0 (0.0) | 0 (0.0) |
| Sepsis | 16 (6.8) | 0 (0.0) | 1 (2.9) | 0 (0.0) | 15 (9.1) | 0 (0.0) | 0 (0.0) |
| Acute bronchitis or exacerbation of chronic bronchitis | 3 (1.3) | 0 (0.0) | 0 (0.0) | 0 (0.0) | 3 (1.8) | 0 (0.0) | 0 (0.0) |
| Gastrointestinal infection | 3 (1.3) | 0 (0.0) | 1 (2.9) | 0 (0.0) | 2 (1.2) | 0 (0.0) | 0 (0.0) |
| Bacteraemia | 2 (0.9) | 0 (0.0) | 2 (5.7) | 0 (0.0) | 0 (0.0) | 0 (0.0) | 0 (0.0) |
| Central nervous system infection | 2 (0.9) | 1 (3.1) | 1 (2.9) | 0 (0.0) | 0 (0.0) | 0 (0.0) | 0 (0.0) |
| Lower urinary tract infection (cystitis) | 2 (0.9) | 0 (0.0) | 1 (2.9) | 0 (0.0) | 1 (0.6) | 0 (0.0) | 0 (0.0) |
| Ear, nose, throat infection ^d^ | 2 (0.9) | 0 (0.0) | 0 (0.0) | 0 (0.0) | 2 (1.2) | 0 (0.0) | 0 (0.0) |
| Febrile neutropenia | 2 (0.9) | 0 (0.0) | 0 (0.0) | 0 (0.0) | 2 (1.2) | 0 (0.0) | 0 (0.0) |
| Bone and joint infection ^e^ | 1 (0.4) | 0 (0.0) | 0 (0.0) | 0 (0.0) | 1 (0.6) | 0 (0.0) | 0 (0.0) |
| Pyrexia of unknown origin | 1 (0.4) | 0 (0.0) | 0 (0.0) | 0 (0.0) | 1 (0.6) | 0 (0.0) | 0 (0.0) |
| Pyrexia of unknown origin in non-neutropenic haemato-oncology patients | 1 (0.4) | 0 (0.0) | 0 (0.0) | 0 (0.0) | 1 (0.6) | 0 (0.0) | 0 (0.0) |

The table lists the 15 most common reasons to prescribe at least one antibiotic for systemic use (J01) by type of hospital-acquired infection. Data are expressed as numbers (percentage) and ranked by frequency. Patients recorded with more than one diagnosis were counted by number of diagnoses. Diagnosis were coded based on the GLOBAL-PPS 2018 Diagnostic Code List.

^a^ Including cellulitis, wound including surgical site infections, deep soft tissue not involving bone (e.g. infected pressure or diabetic ulcers, abscess).

^b^ Including hepatobiliary, intra-abdominal abscess, etc.

^c^ Including catheter related urinary tract infection, pyelonephritis.

^d^ Including mouth, sinuses, larynx.

^e^ Including septic arthritis, including prosthetic joint, osteomyelitis.

^f^ Post-operative surgical site infection (within 30 days of surgery OR; 1 year after implant surgery) (HAI 1)

^g^ Intervention related infections including Catheter-related Blood Stream Infection, Ventilator Associated Pneumonia and Catheter-related Urinary Tract Infection (HAI 2)

^h^ *C. difficile* associated diarrhoea (>48 h post-admission or <30 days after discharge from previous admission episode (HAI 3)

^i^ Other hospital acquired infection (includes Hospital Acquired Pneumonia, etc.) (HAI 4)

^j^ Infection present on admission from another hospital (patient with infection from another hospital) (HAI 5)

^k^ Infection present on admission from long-term care facility or nursing home (HAI 6)

Abbreviations: HAI, Hospital acquired infection; HAP, hospital acquired pneumonia.

**Table S8. Systemic antibiotic use by AWaRe classification**

| **Antibiotic (J01)** | **Total**  **(n=1,273)** | **Access**  **(n=356)** | **Watch**  **(n=858)** | **Reserve**  **(n=31)** | **Unclassified**  **(n=28)** |
| --- | --- | --- | --- | --- | --- |
| Ceftriaxone | 341 (26.8) | 0 (0.0) | 341 (39.7) | 0 (0.0) | 0 (0.0) |
| Levofloxacin | 136 (10.7) | 0 (0.0) | 136 (15.9) | 0 (0.0) | 0 (0.0) |
| Metronidazole | 90 (7.1) | 90 (25.3) | 0 (0.0) | 0 (0.0) | 0 (0.0) |
| Meropenem | 82 (6.4) | 0 (0.0) | 82 (9.6) | 0 (0.0) | 0 (0.0) |
| Cefotaxime | 71 (5.6) | 0 (0.0) | 71 (8.3) | 0 (0.0) | 0 (0.0) |
| Cefoperazone | 52 (4.1) | 0 (0.0) | 52 (6.1) | 0 (0.0) | 0 (0.0) |
| Cefixime | 51 (4.0) | 0 (0.0) | 51 (5.9) | 0 (0.0) | 0 (0.0) |
| Ampicillin sulbactam | 44 (3.5) | 44 (12.4) | 0 (0.0) | 0 (0.0) | 0 (0.0) |
| Gentamicin | 42 (3.3) | 42 (11.8) | 0 (0.0) | 0 (0.0) | 0 (0.0) |
| Amikacin | 32 (2.5) | 32 (9.0) | 0 (0.0) | 0 (0.0) | 0 (0.0) |
| Amoxicillin + clavulanic acid | 31 (2.4) | 31 (8.7) | 0 (0.0) | 0 (0.0) | 0 (0.0) |
| Cefoperazone sulbactam | 28 (2.2) | 0 (0.0) | 0 (0.0) | 0 (0.0) | 28 (100.0) |
| Co-trimoxazole | 28 (2.2) | 28 (7.9) | 0 (0.0) | 0 (0.0) | 0 (0.0) |
| Ceftazidime | 21 (1.6) | 0 (0.0) | 21 (2.4) | 0 (0.0) | 0 (0.0) |
| Ciprofloxacin | 21 (1.6) | 0 (0.0) | 21 (2.4) | 0 (0.0) | 0 (0.0) |
| Cefazolin | 18 (1.4) | 18 (5.1) | 0 (0.0) | 0 (0.0) | 0 (0.0) |
| Ampicillin | 17 (1.3) | 17 (4.8) | 0 (0.0) | 0 (0.0) | 0 (0.0) |
| Amoxicillin | 16 (1.3) | 16 (4.5) | 0 (0.0) | 0 (0.0) | 0 (0.0) |
| Azithromycin | 16 (1.3) | 0 (0.0) | 16 (1.9) | 0 (0.0) | 0 (0.0) |
| Clindamycin | 16 (1.3) | 16 (4.5) | 0 (0.0) | 0 (0.0) | 0 (0.0) |
| Fosfomycin | 15 (1.2) | 0 (0.0) | 0 (0.0) | 15 (48.4) | 0 (0.0) |
| Moxifloxacin | 13 (1.0) | 0 (0.0) | 13 (1.5) | 0 (0.0) | 0 (0.0) |
| Tigecycline | 13 (1.0) | 0 (0.0) | 0 (0.0) | 13 (41.9) | 0 (0.0) |
| Streptomycin | 10 (0.8) | 0 (0.0) | 10 (1.2) | 0 (0.0) | 0 (0.0) |
| Cefadroxil | 9 (0.7) | 9 (2.5) | 0 (0.0) | 0 (0.0) | 0 (0.0) |
| Imipenem and cilastatin | 7 (0.5) | 0 (0.0) | 7 (0.8) | 0 (0.0) | 0 (0.0) |
| Piperacilin tazobactam | 7 (0.5) | 0 (0.0) | 7 (0.8) | 0 (0.0) | 0 (0.0) |
| Cefepime | 6 (0.5) | 0 (0.0) | 6 (0.7) | 0 (0.0) | 0 (0.0) |
| Doripenem | 5 (0.4) | 0 (0.0) | 5 (0.6) | 0 (0.0) | 0 (0.0) |
| Erythromycin | 5 (0.4) | 0 (0.0) | 5 (0.6) | 0 (0.0) | 0 (0.0) |
| Sultamicillin | 5 (0.4) | 5 (1.4) | 0 (0.0) | 0 (0.0) | 0 (0.0) |
| Clarithromycin | 4 (0.3) | 0 (0.0) | 4 (0.5) | 0 (0.0) | 0 (0.0) |
| Cefuroxime | 3 (0.2) | 0 (0.0) | 3 (0.3) | 0 (0.0) | 0 (0.0) |
| Chloramphenicol | 3 (0.2) | 3 (0.8) | 0 (0.0) | 0 (0.0) | 0 (0.0) |
| Vancomycin | 3 (0.2) | 0 (0.0) | 3 (0.3) | 0 (0.0) | 0 (0.0) |
| Penicillin procaine | 2 (0.2) | 2 (0.6) | 0 (0.0) | 0 (0.0) | 0 (0.0) |
| Cefditoren pivoxil | 1 (0.1) | 0 (0.0) | 1 (0.1) | 0 (0.0) | 0 (0.0) |
| Colistin | 1 (0.1) | 0 (0.0) | 0 (0.0) | 1 (3.2) | 0 (0.0) |
| Doxycycline | 1 (0.1) | 1 (0.3) | 0 (0.0) | 0 (0.0) | 0 (0.0) |
| Gentamicin | 1 (0.1) | 1 (0.3) | 0 (0.0) | 0 (0.0) | 0 (0.0) |
| Levofloxacin | 1 (0.1) | 0 (0.0) | 1 (0.1) | 0 (0.0) | 0 (0.0) |
| Linezolid | 1 (0.1) | 0 (0.0) | 0 (0.0) | 1 (3.2) | 0 (0.0) |
| Metronidazole | 1 (0.1) | 1 (0.3) | 0 (0.0) | 0 (0.0) | 0 (0.0) |
| Netilmicin | 1 (0.1) | 0 (0.0) | 1 (0.1) | 0 (0.0) | 0 (0.0) |
| Ofloxacin | 1 (0.1) | 0 (0.0) | 1 (0.1) | 0 (0.0) | 0 (0.0) |
| Polymyxin B | 1 (0.1) | 0 (0.0) | 0 (0.0) | 1 (3.2) | 0 (0.0) |

**Table S9. Quality indicators by ward type**

| **Quality Indicators** | **Total**  **(n=1,273)** | **Medical Ward**  **(n=404)** | **Surgical Ward**  **(n=229)** | **Mixed Ward**  **(n=484)** | **ICU**  **(n=156)** |
| --- | --- | --- | --- | --- | --- |
| Reason documented | 808 (63.5) | 226 (55.9) | 150 (65.5) | 314 (64.9) | 118 (75.6) |
| Stop/review date | 194 (15.2) | 50 (12.4) | 22 (9.6) | 66 (13.6) | 56 (35.9) |
| Treatment duration documented | 125 (9.8) | 38 (9.4) | 24 (10.5) | 49 (10.1) | 14 (9.0) |
| Guideline compliance |  |  |  |  |  |
| Yes | 478 (37.5) | 160 (39.6) | 71 (31.0) | 188 (38.8) | 59 (37.8) |
| No | 378 (29.7) | 90 (22.3) | 77 (33.6) | 148 (30.6) | 63 (40.4) |
| Not Assessable | 358 (28.1) | 117 (29.0) | 77 (33.6) | 131 (27.1) | 33 (21.2) |
| Indication Unknown | 59 (4.6) | 37 (9.2) | 4 (1.7) | 17 (3.5) | 1 (0.6) |
| **Treatment type** |  |  |  |  |  |
| Targeted | 107 (8.4) | 21 (5.2) | 17 (7.4) | 38 (7.9) | 31 (19.9) |
| Empirical | 1,166 (91.6) | 383 (94.8) | 212 (92.6) | 446 (92.1) | 125 (80.1) |
| Route of administration |  |  |  |  |  |
| Parenteral (IV) | 1084 (85.2) | 330 (81.7) | 189 (82.5) | 413 (85.3) | 152 (97.4) |
| Oral | 183 (14.4) | 69 (17.1) | 40 (17.5) | 70 (14.5) | 4 (2.6) |
| IV-oral switch | 48 (26.2) | 7 (10.1) | 21 (52.5) | 20 (28.6) | 0 (0.0) |
| Other | 6 (0.5) | 5 (1.2) | 0 (0.0) | 1 (0.2) | 0 (0.0) |

**Table S10. Bacterial culture sample taken by diagnosis/reason for therapeutic antibiotic use**

| **Diagnosis** | **Total diagnosis^i^**  **(n=777)** | **Total diagnosis with culture**  **(n=344)** | **Blood culture**  **(n=198)** | **Urine culture**  **(n=81)** | **Wound culture**  **(n=37)** | **Sputum culture**  **(n=123)** | **Sterile fluid culture**  **(n=22)** | **Other culture**  **(n=97)** |
| --- | --- | --- | --- | --- | --- | --- | --- | --- |
| Pneumonia or lower respiratory tract infection | 353 (45.4) | 154 (44.8) | 88 (44.4) | 31 (38.3) | 3 (8.1) | 95 (77.2) | 8 (36.4) | 27 (27.8) |
| Skin and soft tissue infections^a^ | 106 (13.6) | 55 (16.0) | 17 (8.6) | 4 (4.9) | 30 (81.1) | 6 (4.9) | 0 (0.0) | 37 (38.1) |
| Gastrointestinal infections | 69 (8.9) | 9 (2.6) | 7 (3.5) | 5 (6.2) | 0 (0.0) | 2 (1.6) | 1 (4.5) | 3 (3.1) |
| Intra-abdominal infections^c^ | 57 (7.3) | 38 (11.0) | 26 (13.1) | 8 (9.9) | 2 (5.4) | 8 (6.5) | 6 (27.3) | 8 (8.2) |
| Sepsis | 46 (5.9) | 24 (7.0) | 23 (11.6) | 4 (4.9) | 0 (0.0) | 5 (4.1) | 1 (4.5) | 5 (5.2) |
| Ear, nose, throat infections^d^ | 30 (3.9) | 10 (2.9) | 1 (0.5) | 1 (1.2) | 1 (2.7) | 0 (0.0) | 0 (0.0) | 9 (9.3) |
| Upper urinary tract infections^e^ | 29 (3.7) | 22 (6.4) | 17 (8.6) | 21 (25.9) | 0 (0.0) | 2 (1.6) | 2 (9.1) | 2 (2.1) |
| Lower urinary tract infections (cystitis) | 19 (2.4) | 5 (1.5) | 2 (1.0) | 3 (3.7) | 0 (0.0) | 2 (1.6) | 0 (0.0) | 1 (1.0) |
| Bone or joint infections^f^ | 13 (1.7) | 5 (1.5) | 1 (0.5) | 0 (0.0) | 1 (2.7) | 1 (0.8) | 2 (9.1) | 3 (3.1) |
| Central nervous system infections | 13 (1.7) | 5 (1.5) | 2 (1.0) | 0 (0.0) | 0 (0.0) | 0 (0.0) | 2 (9.1) | 1 (1.0) |
| Upper respiratory tract viral infections^g^ | 9 (1.2) | 1 (0.3) | 0 (0.0) | 0 (0.0) | 0 (0.0) | 1 (0.8) | 0 (0.0) | 0 (0.0) |
| Cardiovascular system infections | 7 (0.9) | 5 (1.5) | 5 (2.5) | 2 (2.5) | 0 (0.0) | 0 (0.0) | 0 (0.0) | 0 (0.0) |
| Febrile neutropenia | 6 (0.8) | 4 (1.2) | 4 (2.0) | 1 (1.2) | 0 (0.0) | 0 (0.0) | 0 (0.0) | 0 (0.0) |
| Acute bronchitis or exacerbations of chronic bronchitis | 4 (0.5) | 1 (0.3) | 0 (0.0) | 0 (0.0) | 0 (0.0) | 1 (0.8) | 0 (0.0) | 1 (1.0) |
| Other diagnosis^h^ | 4 (0.5) | 1 (0.3) | 1 (0.5) | 0 (0.0) | 0 (0.0) | 0 (0.0) | 0 (0.0) | 0 (0.0) |

The table lists the 15 most common reasons to treat adult inpatients for CAI or HAI with at least one antibiotic for systemic use (J01). Data are expressed as numbers (percentage) and ranked by frequency. Patients recorded with more than one diagnosis were counted by number of diagnoses. Diagnosis were coded based on the GLOBAL-PPS 2018 Diagnostic Code List.

^a^	Including cellulitis, wound including surgical site infections, deep soft tissue not involving bone (e.g. infected pressure or diabetic ulcers, abscess).

^b^	Including prophylaxis for surgery of the gastrointestinal tract, liver, or biliary tree, and prophylaxis in patients with neutropenia or hepatic failure.

^c^	Including hepatobiliary, intra-abdominal abscess, etc.

^d^	Including mouth, sinuses, larynx.

^e^	Including catheter related urinary tract infection, pyelonephritis.

^f.^	Including septic arthritis, including prosthetic joint, osteomyelitis.

^g^	Including influenza but not ear, nose, throat infections.

^h^	Antibiotic prescribed with documentation for which there is no above diagnosis group.

^i^	Total number of diagnosis/reasons for systemic antibiotic use for treatment of community- or hospital-acquired infection

Abbreviations: CAI, community acquired infection; HAI, hospital acquired infection
